# Integrated Proteomic and Network Analysis Reveals Dysregulated Pathways and Candidate Proteins in Multiple Myeloma Progression

**DOI:** 10.64898/2026.05.21.26353799

**Authors:** Foteini Paradeisi, Charis Gonidaki, Aggeliki Tserga, Julie Courraud, Panagiotis Bakouros, Paraskevi Karousi, Ioannis V. Kostopoulos, Theodoros Margelos, Eirini Goula, Clara Stegehuis, Janusz M. Meylahn, Anastasia Martzakli, Christine-Ivy Liacos, Meletios A. Dimopoulos, Ourania Tsitsilonis, Antonia Vlahou, Jerome Zoidakis, Efstathios Kastritis

**Author notes:** **Corresponding Author:** Jerome Zoidakis.

## Abstract

**Background:** Multiple myeloma (MM) remains incurable despite therapeutic advances, reflecting limited understanding of the molecular mechanisms underlying disease initiation and progression. MM develops through asymptomatic precursor stages, monoclonal gammopathy of undetermined significance (MGUS) and smouldering multiple myeloma (SMM). This study aimed to investigate protein changes associated with disease progression and, through a further integrative approach, to highlight molecular changes of potential predictive and/or therapeutic value.

**Methods:** We performed a comparative proteomic analysis of 94 bone marrow–derived CD138⁺-selected plasma cell samples (29 MGUS, 20 SMM, and 45 MM) using LC–MS/MS. Differential protein abundance was assessed using pairwise Mann–Whitney *U* tests between groups, with Benjamini–Hochberg correction. Pathway enrichment, protein–protein interaction, and co-expression network analyses were also conducted. Selected proteins were further evaluated using public transcriptomic datasets and experimentally validated in independent samples by flow cytometry and enzyme-linked immunosorbent assay (ELISA).

**Results:** Following data processing, proteomic analysis identified 6,203 proteins. Pairwise comparisons revealed significant proteomic differences across disease stages, with 370 differentially abundant proteins exhibiting monotonic changes during disease progression. Pathway analysis showed that monotonically upregulated proteins were mainly associated with gene expression and cell proliferation, whereas downregulated proteins were linked to immune-related processes. Further co-expression network analysis, combined with criteria including detection frequency, biological relevance, and translational potential, highlighted a group of prioritised proteins. Representative examples include nucleolin (NCL) and U3 small nucleolar ribonucleoprotein IMP3 (IMP3), involved in nucleolar organisation, ribosome biogenesis and rRNA processing, as well as the immune-associated lactotransferrin (LTF) and serine protease cathepsin G (CTSG). Transcriptomic support and independent experimental validation by flow cytometry and ELISA confirmed the relevance of selected candidates.

**Conclusions:** Taken together, our findings highlight coordinated changes in immune regulation, RNA processing and ribosome biogenesis during MM progression and identify candidate proteins and their networks, including the emerging pharmacologically tractable target NCL and the underexplored IMP3 of potential therapeutic relevance, opening new avenues for further investigation.

## 1. BACKGROUND

Multiple myeloma (MM) is a haematological malignancy characterised by the clonal proliferation and accumulation of plasma cells (PCs) in the bone marrow (BM), which secrete a monoclonal immunoglobulin detectable in the blood and/or urine. It is the second most common blood cancer, accounting for approximately 1% of all cancers and 10-15% of haematological malignancies [1–3] and arises from two asymptomatic precursor stages: monoclonal gammopathy of undetermined significance (MGUS) and smouldering multiple myeloma (SMM) [4].

Despite important advances in diagnosis and treatment (e.g. immunomodulatory drugs, proteasome inhibitors, corticosteroids, and monoclonal and bispecific antibodies), MM remains incurable due to frequent relapses and the development of drug resistance [5,6]. Moreover, a major clinical challenge remains in predicting which patients with precursor conditions (MGUS or SMM) will eventually progress to symptomatic disease and could benefit from early therapeutic intervention [7]. Addressing these challenges requires a deeper understanding of the molecular pathways underlying myelomagenesis and disease progression.

In this context, the application of state-of-the-art omics technologies provides a powerful approach for elucidating key regulators associated with disease onset and evolution. To date, most studies investigating MM development have primarily focused on genomic and transcriptomic profiling [8].

Specifically, large-scale genomic analyses have linked disease progression to the activation of oncogenic drivers such as *MYC* and *RAS*, chromosomal instability, and copy number alterations [9]. Bulk transcriptomic studies further showed that progression to MM involves metabolic reprogramming together with alterations in antigen presentation [10–15].

Single-cell approaches have provided additional insights into the cellular heterogeneity underlying disease progression. In particular, single-cell RNA sequencing studies across MGUS to MM progression have revealed reduced immune-related programs (e.g. antigen receptor signaling), loss of normal PC identity, decreased expression of apoptosis-associated genes, heterogeneous activation of oncogenic pathways, and proteasome upregulation during evolution to overt MM [16–19]. While these studies provide valuable information, they largely fail to capture the proteomic component and its direct link to phenotype [20]. Investigation at the protein level remains limited, with only a small number of single-cell proteomics studies available, demonstrating reduced T-cell representation in the BM microenvironment in MM, consistent with progressive immune suppression [21], as well as increased activation of proliferative and survival pathways and upregulation of adhesion-related molecules in symptomatic disease [22].

In this regard, untargeted mass spectrometry-based proteomics via liquid chromatography-tandem mass spectrometry (LC-MS/MS) enables the direct investigation of proteome-wide alterations [20]. Data generated by these efforts have provided insights into disease-associated processes such as extracellular matrix remodelling, angiogenesis, and immune evasion, highlighting the interplay between the BM microenvironment and MM development [20,23,24]. However, limited studies have examined proteomic alterations across the disease continuum from MGUS and SMM to symptomatic MM [25].

The aim of this study was the identification of altered proteins and dysregulated molecular pathways associated with disease onset and progression. To this end, we compared the proteomes of BM-derived CD138⁺-selected PCs from patients spanning the disease spectrum from precursor conditions (MGUS, SMM) to symptomatic MM. Proteomic analysis was performed using LC-MS/MS and specifically the high-resolution Data Independent Acquisition (DIA) method [26], followed by statistical and bioinformatics analyses. Key findings were further evaluated using publicly available transcriptomic datasets and validated in independent clinical samples using complementary immunophenotypic and protein quantification assays (**Graphical abstract**).

## 2. METHODS

### 2.1. Patient Cohort

We performed proteomic analysis on BM-derived CD138⁺ PC samples from 29 MGUS, 20 SMM, and 45 newly diagnosed MM (NDMM) patients. All patients had both a BM trephine biopsy and a BM aspirate collected at baseline (prior to any treatment in patients with symptomatic disease), and sample classification was based on the International Myeloma Working Group (IMWG) criteria [1]. All patients provided written informed consent for the collection of biological material and clinical data, as well as for their analysis, following approval by the Ethics/Scientific Committee of Alexandra Hospital (No 111368/25.10.2022).

### 2.2. Isolation of CD138+ Cells from Bone Marrow Aspirates

BM aspirates collected in EDTA tubes were processed for the isolation of CD138+ cells using the AutoMACS Pro Separator (Miltenyi Biotec) as previously described [27]. Briefly, samples were directly labeled with anti-CD138 microbeads and separated according to the manufacturer’s instructions. In selected cases with low cell yield or limited sample volume, mononuclear cells were first enriched by density gradient centrifugation prior to CD138 labeling.

### 2.3. Sample Preparation for Proteomics

Pellets from BM-derived CD138⁺ PCs were subjected to nucleic acid extraction using TRI Reagent® (Molecular Research Center Inc., Cincinnati, OH, USA), according to the manufacturer’s protocol, and the organic (phenol) phase of each sample was retained for subsequent proteomic analysis. These extracts were processed using an optimised, adapted Sample Preparation by Easy Extraction and Digestion (SPEED) protocol as described by Karousi et al. [27].

In brief, proteins were precipitated by incubation with acetone at 4 °C for 4 h, followed by centrifugation at 18,200 RCF for 5 min at 4 °C. The protein pellets were denatured using trifluoroacetic acid (TFA), and an amount equivalent to 30,000 cells was transferred to a new tube and neutralised with 6-8 volumes of 2 M Tris base. Reduction and alkylation were conducted by adding a solution of Tris(2-carboxyethyl)phosphine hydrochloride (TCEP.HCl, 100 mM) and 2-chloroacetamide (CAA, 400 mM) at 10% (v/v), followed by vortexing and heating at 95 °C for 5 min with shaking at 400 rpm. Water was subsequently added to reduce the Tris base concentration to below 0.5 M, ensuring a pH of 8-9. Proteolytic digestion was carried out overnight at 37 °C using 40 ng of MS-grade Trypsin Gold (Promega), with shaking at 400 rpm. Digestion was terminated and peptides were acidified by adding approximately 2% of TFA (v/v).

Peptides were purified and dried as previously described [27] and reconstituted in 10 μL of 0.5% acetic acid in H₂O per 10,000 cells shortly prior to LC-MS/MS analysis.

### 2.4. LC-MS/MS Analysis

For LC-MS/MS analysis, following the method of Karousi et al. [27], peptides were injected onto an analytical column (PepSep #1893477, 25 cm × 75 μm, 1.9 μm beads, C18 ReproSil AQ; Bruker GmbH, Mannheim, Germany), and gradient elution was performed using a nanoElute® 2 system (Bruker Daltonics GmbH) over a 30-min run at a flow rate of 300 nL/min. Separated peptides were analysed on a timsTOF fleX mass spectrometer (Bruker Daltonics GmbH). Mass spectra were acquired in library-free data-independent acquisition with parallel accumulation–serial fragmentation (dia-PASEF) mode across an optimised mass range of *m/z* 300–1300.

### 2.5. MS Data Processing

Raw LC-MS/MS files were subsequently processed using FragPipe v22.0 [28] with DIA-NN v1.9.0 [29] in library-free mode. DIA-Tracer was utilised to improve feature alignment and quantification across runs [30]. Search parameters allowed up to four missed cleavages, with peptide lengths of 5-30 amino acids and peptide masses between 300 and 5,000 Da. A false discovery rate (FDR) of 1% was applied at the precursor, peptide, and protein levels. Database searches were performed against the human reference proteome (Uniprot reference proteome UP000005640, one protein sequence per gene [31]), permitting up to two variable modifications per peptide. These included oxidation of methionine residues and N-terminal methionine excision, with carbamidomethylation of cysteine residues specified as a fixed modification.

Thereafter, the resulting protein intensity data were processed using the R programming language (v4.4.0 [32]). For each sample, protein intensities were normalised by parts per million (ppm): [(protein intensity/total intensity in the sample) × 10⁶], calculated after excluding highly abundant proteins of putative plasma origin (albumin, haemoglobins, and haptoglobin), to better reflect the cellular proteome of CD138⁺ PCs. Proteins detected in at least 50% of samples in at least one disease stage were retained for further analysis. Given the high sensitivity and reproducibility of DIA-based proteomics [26], missing values, commonly associated with low-intensity signals near the detection limit, were imputed with the lowest observed intensity. Protein intensities were then log₂-transformed. Principal component analysis (PCA) was performed in R using the prcomp function, and PCA plots were generated using the ggplot2 (v4.0.1) package.

### 2.6. Statistical Analysis of the Proteomic Data

Differential protein abundance between disease stages was assessed in R (v4.4.0) using the non-parametric Mann–Whitney *U* test for pairwise comparisons, with all SMM samples considered as a single group, irrespective of risk stratification, to increase statistical power. For each comparison, both raw and adjusted p-values (Benjamini-Hochberg method [33]) were calculated.

Proteins were considered differentially abundant (DAPs) in a given comparison if they exhibited an adjusted p-value < 0.05 and a fold change (FC) ≥ 1.5 for upregulated proteins or ≤ 0.67 for downregulated proteins. Monotonic DAPs were defined as proteins that showed differential abundance in the pairwise comparisons between MM vs MGUS and MM vs SMM and also exhibited a monotonic increase (MGUS < SMM < MM) or decrease (MGUS > SMM > MM) in mean abundance across stages. Volcano plots were generated using the ggplot2 (v4.0.1) package, and heatmaps were generated using the ComplexHeatmap (v2.20.0) package.

### 2.7. Bioinformatic Analyses

#### 2.7.1. Pathway Enrichment Analysis

Pathway enrichment analysis was performed separately for monotonically upregulated and downregulated DAPs, using the clusterProfiler (v4.14.3), ReactomePA (v1.50.0), and msigdbr (v10.0.0) packages in R (v4.4.1) [34–36]. Enrichment analysis for Gene Ontology Biological Process (GO-BP) terms, Reactome pathways, and Hallmark gene sets was performed with the compareCluster function with default settings. GO-BP enrichment was conducted using the enrichGO function, Reactome pathway enrichment with enrichPathway, and Hallmark gene-set enrichment with the enricher function using Hallmark gene sets from the Molecular Signatures Database (MSigDB) [36]. Enrichment results were visualised using custom dot plots generated with ggplot2 (v3.5.1). Pathways with an adjusted p-value <0.05, corrected for multiple testing using the Benjamini-Hochberg method [33], were considered statistically significant. Selected pathways are presented based on biological relevance.

#### 2.7.2. Protein-Protein Interaction Analysis

Protein-protein interaction (PPI) data were obtained from STRING (v12.0) [37] using the STRING application within Cytoscape (v3.10.3) [38]. Neighbouring proteins in the PPI network represent proteins with known or predicted physical or functional associations, which can indicate participation in shared molecular processes or pathways [39]. High-confidence interactions were kept by applying a STRING combined confidence score threshold of 0.9. Network visualisation was performed using a Fruchterman-Reingold layout and visualised in R (v4.4.1) with ggplot2 (v3.5.1).

Community structure within the resulting PPI network was identified using the cluster_walktrap function from the igraph package (v2.1.1) [40]. The “steps” parameter of the algorithm was evaluated across a range of values (2-10). For each value, clustering stability, modularity, and the number of detected communities were computed. The final choice (steps = 7) was selected as it maximised clustering stability and showed high modularity (> 0.43), while avoiding parameter values that produced trivial partitions, such as a single large cluster or many very small clusters of one or two proteins. Stability was assessed by repeated edge perturbation (5% random edge removal, 50 replicates) and quantified using the adjusted Rand index (ARI), as implemented in the mclust package (v6.1.1) [41]. The identified PPI clusters were functionally characterised based on pathway enrichment analysis using the Reactome database [42].

#### 2.7.3. Co-Expression Network Analysis

Co-expression networks were constructed separately for each disease stage (MGUS, SMM, and MM) using the processed protein expression data. Neighbouring proteins in co-expression networks represent proteins with coordinated abundance patterns across samples, which may reflect involvement in related biological processes or functional modules [43]. The networks were created using R (v4.4.1) by applying the GENIE3 algorithm (v1.28.0) [44] with 100 trees. GENIE3 produces directed weighted association scores between protein pairs. These were converted into undirected weighted networks by averaging edge weights in both directions and self-loops were removed, yielding one undirected weighted co-expression network per disease stage. Proteins detected in <50% of samples in a given stage were kept as nodes but forced to be isolated in that stage (edges set to zero) to reduce low-coverage artefacts.

To obtain a sparse and interpretable network representation, only high-confidence edges were retained by applying a quantile-based threshold corresponding to the top 0.5% strongest edges (99.5th percentile). The threshold was selected in a data-driven way in order to satisfy multiple structural criteria across all stages, including: (i) network density within a predefined mean-degree range (25-35), (ii) preservation of a sufficiently large connected component (≥30% of all nodes), (iii) retention of a minimum fraction of active (non-isolated) proteins (≥70%), and (iv) stability of the top-ranked proteins under small changes in the threshold, assessed by comparing the overlap of the top 100 highest-degree proteins identified at adjacent quantile values using the Jaccard index (≥60% overlap).

Following thresholding, the weighted adjacency matrices were binarised by assigning a value of 1 to edges with weights above the selected threshold and 0 otherwise, resulting in a binary co-expression network for each disease stage. For each protein, degree (i.e., connectivity) was defined as the number of connected neighbours in the corresponding binary network and degree values for all three stages were reported. To quantify stage-dependent changes, pairwise differences in degree between stages were calculated, with a particular focus on the MM vs MGUS comparison (Δ = degree_MM − degree_MGUS), representing respectively the latest and earliest disease states. In addition to changes in connectivity, similarity of protein neighbourhoods across stages was assessed using the overlap coefficient, defined as the fraction of shared neighbours normalised by the smaller of the two neighbourhood sizes. All co-expression subnetworks were visualised in R (v4.4.1) using igraph (v2.1.1) and ggplot2 (v3.5.1) with a Fruchterman-Reingold force-directed layout.

### 2.8. Validation of Proteomic Findings

#### 2.8.1. Validation Using Public Transcriptomic Data

Publicly available human transcriptomic studies related to MGUS, SMM, and MM were identified through PubMed [45], ArrayExpress [46] and the Gene Expression Omnibus [47] repositories. Searches were performed using combinations of the terms “MM”, “MGUS”, “SMM”, “transcriptom*”, and “CD138”. Additional disease-related terms were also queried, including “multiple myeloma”, “monoclonal gammopathy of undetermined significance”, and “smouldering multiple myeloma”.

Based on information provided in abstracts and accompanying data, three studies [10,11,48] were identified that reported differential expression results derived from CD138⁺ PCs across disease-stage comparisons involving MGUS, SMM, MM, and control donors. Differential expression results were used as reported by the respective authors.

#### 2.8.2. Validation Using ELISA and Flow Cytometry

To quantify secreted lactoferrin (lactotransferrin, LTF) levels and cellular cathepsin G (CTSG) expression, we used enzyme-linked immunosorbent assay (ELISA) and multi-color flow cytometry (MFC), respectively. Soluble LTF levels were measured in BM plasma samples obtained from 100 patients (30 MGUS, 30 SMM and 40 NDMM). Analysis was performed using the Human LTF/Lactoferrin ELISA Kit (Catalogue #EEL069, Invitrogen, Thermo Fisher Scientific, MA), following the manufacturer’s instructions. Briefly, BM plasma was pre-diluted at a ratio of 1:250 with sample diluent before analysis. The assay utilises a sandwich format with a detection range of 0.625–40 ng/mL and a minimum detectable concentration (sensitivity) of 0.375 ng/mL. The final concentrations were determined by interpolation from a standard curve.

To assess CTSG expression specifically on clonal PCs, flow cytometry was performed using 100 cryopreserved total BM nucleated cell samples (30 MGUS, 30 SMM and 40 NDMM) retrieved from the biobank of the Flow Cytometry Unit, National and Kapodistrian University of Athens. After thawing, cells were first stained with eBioscience™ Fixable Viability Dye eFluor™ 450 (Thermo Fisher Scientific) to exclude dead cells from analysis. Staining for surface markers was performed using a 7-color panel comprising the following fluorophore-conjugated antibodies: CD38-FITC (multi-epitope; BD Biosciences, CA), CD45-PerCP/Cy5.5 (clone HI30; BD Biosciences), CD27-BV510 (clone O323; BioLegend, CA), CD19-PC7 (clone J3-119; Bechman Coulter, CA), CD56-PE (clone C5.9; BD Biosciences), CD81-APC-C750 (clone M38; BD Biosciences), and Cathepsin G-APC (polyclonal; Assaypro, MO). Data acquisition was performed on a Cytek Northern Lights (Cytek Biosciences, CA), and subsequent analysis was conducted using FlowJo software (BD Biosciences). Clonal PCs were identified and gated based on a CD38^+^/CD45^dim^ phenotype combined with the presence of aberrant marker expression [49], through which the surface levels of CTSG were quantified.

Statistical analysis for ELISA and flow cytometry experiments was performed using one-way ANOVA, followed by Tukey’s post hoc test for pairwise comparisons. Statistical significance was defined as p-value < 0.05.

## 3. RESULTS

### 3.1. Patient characteristics

Samples from a total of 94 patients were processed for proteomics analysis, comprising 29 MGUS, 20 SMM, and 45 MM cases. The main demographic and clinical characteristics of the patients are presented in **Table 1**.

**Table 1:** Patients baseline characteristics.

|  |  | **MGUS** | **SMM** | **MM** |
| --- | --- | --- | --- | --- |
| **Sex** | Female | 17 | 11 | 20 |
|  | Male | 12 | 9 | 25 |
| **Age (years)** | Median [min-max] | 68 [41-86] | 67 [48-82] | 68 [42-86] |
| **SMM risk category<sup>a</sup>** | Low | - | 11 | - |
|  | Intermediate | - | 7 | - |
|  | High | - | 1 | - |
|  | Missing | - | 1 | - |
| **Ratio of kappa FLC/lambda FLC<sup>b</sup>** | Median [min-max] | 1.4 [0.1-9.3] | 1.4 [0-34.2] | - |
|  | Missing | 5 | 1 | - |
| **Light chain type** | Kappa | 22 | 10 | 33 |
|  | Lambda | 6 | 10 | 11 |
|  | Missing | 1 | 0 | 0 |
| **Heavy chain type (isotype)** | IgG | 25 | 16 | 29 |
|  | IgA | 2 | 3 | 6 |
|  | IgM | 1 | 0 | 1 |
|  | Missing | 1 | 0 | 0 |
| **International Staging System (ISS) stage for prognosis** | 1 | - | - | 11 |
|  | 2 | - | - | 9 |
|  | 3 | - | - | 21 |
|  | Missing | - | - | 4 |
| **Albumin (g/dL)** | Median [min-max] | 4.6 [3.0-5.1] | 4.4 [3.6-4.9] | 3.7 [2.2-5.2] |
|  | Missing | 1.0 | 0 | 0 |
| **Creatinine (mg/dL)** | Median [min-max] | 0.8 [0.4-4.4] | 0.9 [0.6-1.3] | 1.0 [0.4-9.3] |
|  | Missing | 1 | 0 | 0 |
| **Hemoglobin (g/dL)** | Median [min-max] | 13.2 [4.7-15.6] | 12.3 [10.7-15.5] | 10.5 [4.4-14.8] |
|  | Missing | 0 | 0 | 0 |
| **M-spike (g/dL)** | Median [min-max] | 0.5 [0-2.0] | 1.4 [0-2.8] | 3.0 [0-9.5] |
|  | Missing | 4 | 1 | 3 |
| **Beta-2 Microglobulin (mg/L)** | Median [min-max] | 2.0 [1.1-18.3] | 2.1 [1.2-4.4] | 4.9 [0.2-36.5] |
|  | Missing | 8 | 6 | 0 |
| **Plasma cell % in bone marrow trephine biopsy** | Median [min-max] | 0 [0-10] | 11 [10-60] | 70 [10-100] |
|  | Missing | 3 | 0 | 2 |

Baseline is defined as the period close to the date of diagnosis and before any therapy is initiated. Measurements were performed in blood unless otherwise specified.
<sup>b</sup>FLC: free light chains
<sup>a</sup>SMM risk categories as defined in Mateos et al. 2020 [50]

### 3.2. Proteomic differences across disease stages

Considering only proteins detected in at least 50% of samples within at least one disease stage, the analysis of the 94 BM-derived CD138⁺ PC samples from MGUS, SMM, and MM patients enabled the identification and relative quantification of 6,203 proteins (**Additional file 1**). Imputed missing values accounted for approximately 14% of the filtered dataset. Global principal component analysis (PCA) revealed distinct but slightly overlapping proteomic profiles between MGUS and MM samples, while SMM displayed an intermediate proteomic profile, consistent with its transitional phenotype (**Figure 1A**).

**Figure 1:**
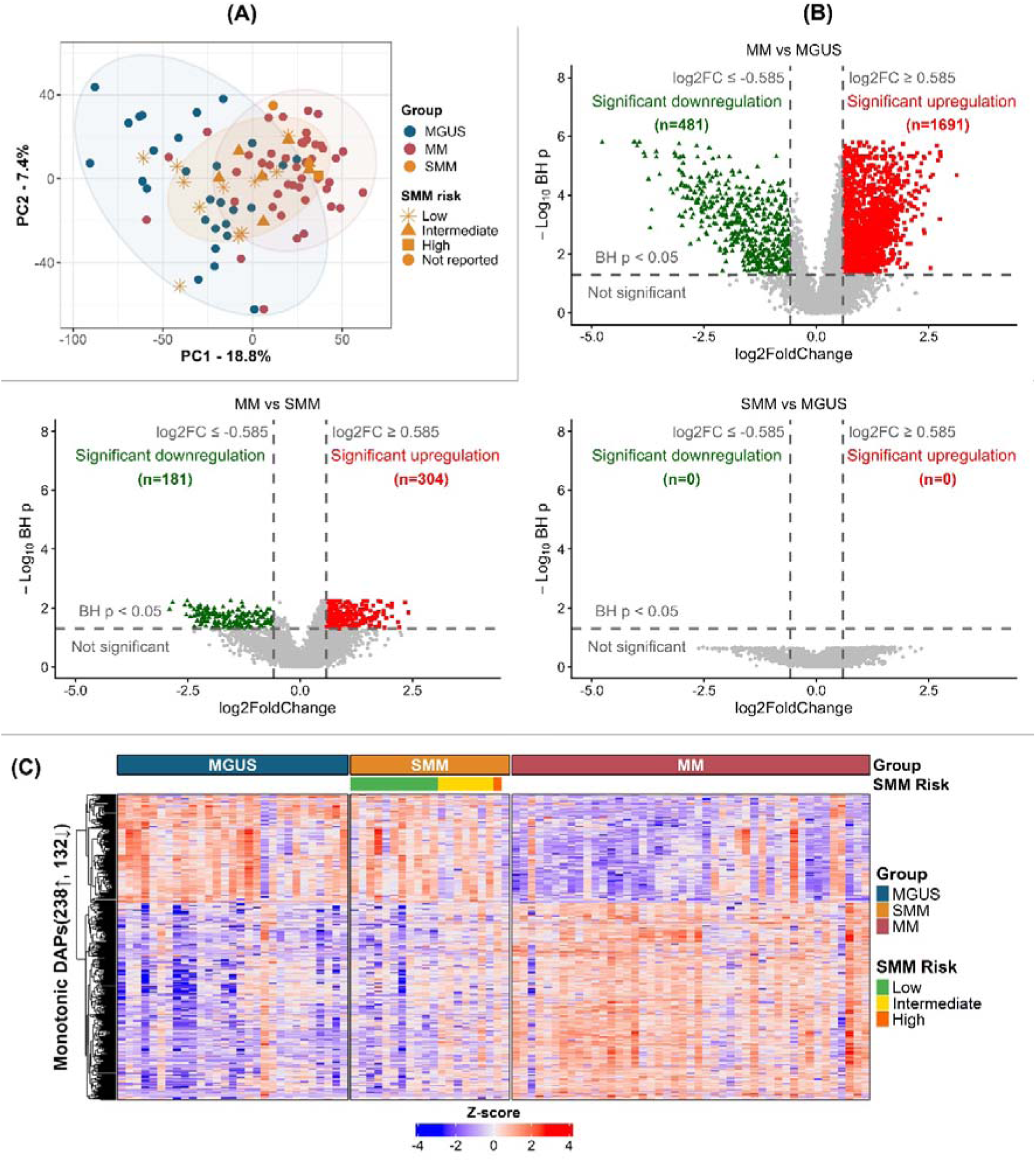
Proteomic differences across disease stages. **(A)** Global PCA plot based on the filtered proteomic dataset (6,203 proteins) depicting the distribution of MGUS, SMM and MM samples based on protein abundance. The x-axis (PC1) and y-axis (PC2) represent the first two principal components, explaining the largest proportion of variance in protein abundance. Each point represents an individual sample and is coloured according to disease stage (MGUS, SMM, MM). The observed separation between MGUS and MM samples, with SMM occupying an intermediate position, reflects stage-associated differences in the global proteomic profile. **(B)** Volcano plots depicting DAPs across pairwise comparisons between disease stages. The x-axis represents log_₂_ fold change and the y-axis −log_₁₀_ Benjamini-Hochberg-adjusted p-values derived from the Mann-Whitney U test. Proteins with adjusted p-value < 0.05 and |log_₂_FC| ≥ 0.585 (corresponding to a fold change ≥1.5 or ≤0.67) were considered significant. **(C)** Heatmap depicting the abundance patterns of the 370 DAPs exhibiting a monotonic trend across disease progression. Columns represent individual samples grouped by disease stage (MGUS, SMM, and MM), and rows represent individual proteins. Protein intensities were standardised using z-score scaling across samples. Colour intensity reflects relative protein abundance following z-score scaling, with red indicating higher and blue indicating lower abundance across samples. **Abbreviations**: BH: Benjamini-Hochberg; DAPs: Differentially Abundant Proteins; log_₂_FC: log_₂_ fold change; PCA: Principal Component Analysis.

Differential protein abundance across disease stages is summarized in **Figure 1B**. Consistent with PCA, no DAPs (adjusted p-value < 0.05; FC ≥ 1.5 or ≤ 0.67) were observed between SMM and MGUS, indicating proteomic overlap between the two groups, in line with the relatively high representation of low-risk SMM cases in our cohort, which exhibit a clinical course similar to MGUS [51]. In contrast, the highest number of DAPs was observed in the comparison between MM and MGUS, reflecting the larger molecular divergence between these clinically distinct stages.

Among the DAPs, 370 were statistically significant in both the MM vs MGUS and MM vs SMM comparisons and exhibited monotonic changes in mean abundance across the three disease stages. These proteins were defined as monotonic DAPs (**Methods 2.6**). Of these, 238 proteins showed a progressive increase (MGUS < SMM < MM), while 132 displayed a progressive decrease (MGUS > SMM > MM) (**Figure 1C**). A complete list of the 370 monotonic DAPs is provided in **Additional file 1**.

### 3.3. Monotonic proteome changes reflect alterations in RNA processing and immune functions

To investigate the biological relevance of the proteins exhibiting monotonic changes in abundance, pathway enrichment analysis was performed for the monotonically up- and downregulated DAPs using the Gene Ontology Biological Process, Reactome databases, and Hallmark gene sets. The results (**Figure 2A**; **Additional file 2**) indicate that monotonically upregulated proteins are mainly associated with pathways related to cell-cycle progression (e.g., E2F targets, G2M checkpoint, DNA replication), ribosome biogenesis, MYC target gene sets, and the unfolded protein response (UPR). In contrast, monotonically downregulated proteins are predominantly related to pathways involved in immune response regulation (e.g. antigen processing and presentation, interferon signaling, and regulation of immune effector processes).

**Figure 2:**
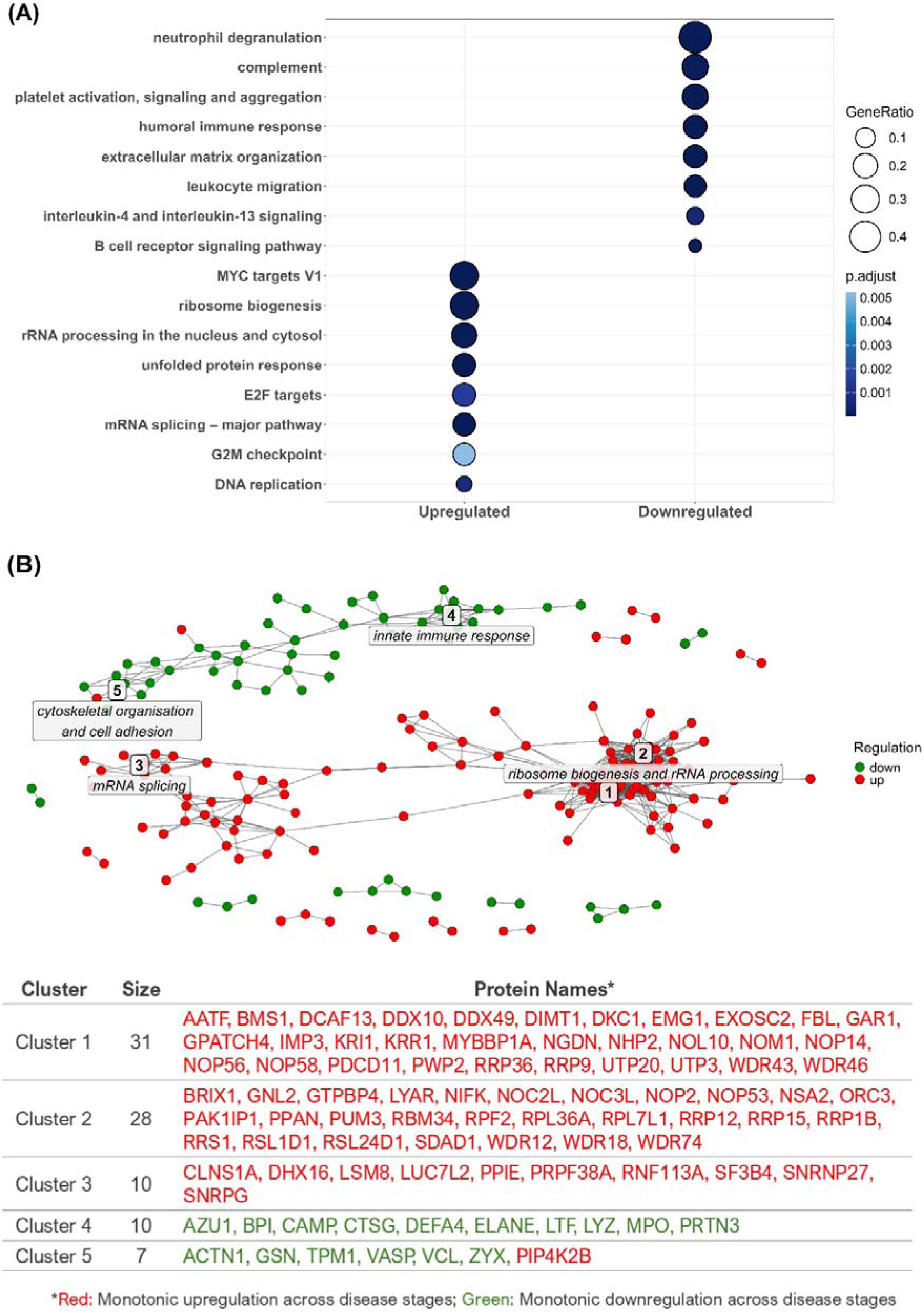
Functional enrichment and PPI network of monotonic DAPs. **(A)** Selected enriched Gene Ontology Biological Process (GO-BP) terms, Reactome pathways and Hallmark gene sets associated with the monotonically up- and downregulated DAPs. Dot size represents the number of proteins associated with each pathway, and dot colour indicates the adjusted p-value. All pathways shown are statistically significant after Benjamini-Hochberg adjustment for multiple testing (adjusted p-value < 0.05)**. (B)** Subgraph of the PPI network restricted to the 370 monotonic DAPs. Protein pairs with high-confidence interactions (STRING confidence score > 0.9) are connected by an edge. Nodes represent proteins, with red and green indicating monotonically upregulated and downregulated DAPs, respectively. The five largest communities (clusters) identified in the network are labelled from 1 to 5 and shown together with descriptive functional titles.

To further explore how these proteins may be interacting at network level, we extracted the PPI subnetwork induced by these 370 monotonic DAPs and examined its community structure. This analysis revealed five major functional clusters. Clusters 1-3 consist exclusively of monotonically upregulated proteins, whereas Clusters 4 and 5 are primarily composed of monotonically downregulated proteins. More specifically, Clusters 1-3 are associated with ribosome biogenesis and ribosomal RNA (rRNA) processing (Clusters 1,2), as well as mRNA splicing (Cluster 3). In contrast, Cluster 4 is associated with the innate immune response, and Cluster 5 is characterised by proteins related to cytoskeletal organisation and cell adhesion. An overview of the PPI clusters is shown in **Figure 2B**, and the degree and corresponding interaction partners for each protein in the entire dataset are reported in **Additional file 3**.

### 3.4. Co-expression network connectivity changes reveal candidate proteins associated with MM progression

To place these findings within a holistic framework and allow the identification of potential central nodes, stage-specific co-expression analysis was further performed. To ensure high coverage, all 6,203 proteins were considered as network nodes in the analysis, resulting in networks of more than 96,000 edges per stage (density = 0.005; mean degree = 31).

Protein degrees were computed for each disease stage, and proteins were ranked according to the absolute change in degree between MM and MGUS (|Δdegree| = |degree_MM − degree_MGUS|), as defined in **Methods 2.7.3**, to capture the largest progression-associated gains or losses in connectivity. Stage-specific connectivity metrics for the monotonic DAPs are provided in **Additional file 4**.

To shortlist the most prominent findings, we focused on the monotonic DAPs falling within the top 30% of proteins exhibiting the largest absolute connectivity changes. This resulted in 174 proteins showing evident shifts in their co-expression patterns as disease progresses (**Additional file 5**). Their degree values in each stage and their corresponding neighbours are reported in **Additional file 4.**

Among these, RNA-binding proteins and ribosome biogenesis factors (e.g., DDX helicases, IMP3, ILF3, NCL) represented a dominant functional category, indicating extensive remodelling of RNA processing and translational machinery during progression to MM. In parallel, proteins involved in DNA replication, nucleotide metabolism, and cell-cycle control (e.g., TK1, TOP2B, DUT, SUMO2) also displayed changes in connectivity, consistent with proliferation-associated alterations in network structure. Proteins involved in innate immunity, including CTSG, CAMP, and LTF, were also represented within this set.

To further prioritise and shortlist proteins with strong biological relevance the following selection criteria were applied:

- High detection frequency of each protein across disease stages (≥90% of samples per stage) or a clear stage-associated detection pattern (e.g. progressive increase or decrease in detection frequency across stages, consistent with corresponding changes in protein abundance);
- Documented expression in the BM tissue and myeloma cell lines, based on publicly available resources (Human Protein Atlas [52,53]);
- Evaluation of prior literature linking the proteins to the BM microenvironment, MM pathogenesis, or broader cancer-associated mechanisms, to contextualise their established or less-characterised roles in MM;
- Translational potential, as indicated by the availability of FDA-approved or investigational agents in clinical development targeting the proteins, where applicable.

Application of these criteria resulted in a focused list of 22 prioritised proteins capturing the principal biological pathways identified above, highlighting established MM-associated factors as well as potentially novel candidates with relevance for biomarker development, therapeutic targeting, or mechanistic investigation in MM (**Table 2**).

**Table 2:** Prioritised Proteins Representing Key Biological Pathways in MM Progression.

| Protein | Biological Function (UniProt [31]) & Literature Evidence | Therapeutic Relevance (DrugBank [54]) | Fold Change<sup>a</sup> |  |  |  | Co-Expression Network Analysis Metrics<sup>b</sup> |  |  |
| --- | --- | --- | --- | --- | --- | --- | --- | --- | --- |
|  |  |  | SMM vs MGUS | MM vs SMM | MM vs MGUS | Trend | Degree MGUS | Degree SMM | Degree MM |
| U3 small nucleolar ribonucleoprotein IMP3 (**IMP3**) | RNA-binding component of the U3 small nucleolar ribonucleoprotein (snoRNP) complex required for early pre-18S rRNA processing during ribosome biogenesis. |  | 1.79 | 1.66 * | 2.97 *** | Monotonically upregulated | 23 | 37 | 92 |
| Interleukin enhancer-binding factor 3 (**ILF3**) | RNA-binding protein regulating circular RNA biogenesis and RNA stability; reported to be upregulated in MM and shown to promote MM cell proliferation via m6A-dependent AKT3 mRNA stabilization [55]. | Sepantronium bromide - YM155 (Clinical-stage, Phase I/II) | 1.10 | 1.50 ** | 1.66 *** | Monotonically upregulated | 28 | 33 | 89 |
| RNA-binding protein 5 (**RBM5**) | RNA-binding spliceosomal protein regulating alternative splicing of multiple mRNAs involved in apoptosis and cell-cycle control; RBM5 knockdown in cell-line models has been reported to impair osteoclast differentiation [56]. |  | 1.28 | 1.50 * | 1.92 ** | Monotonically upregulated | 34 | 45 | 93 |
| Cell growth-regulating nucleolar protein (**LYAR**) | Nucleolar regulator of ribosome biogenesis; described as an oncogenic factor in MM and downregulated following treatment with TAS0612 (an investigational triple kinase inhibitor) in MM cell lines [57]. |  | 1.35 | 1.75 * | 2.36 *** | Monotonically upregulated | 26 | 67 | 81 |
| E3 ubiquitin-protein ligase RNF113A (**RNF113A**) | Spliceosome-associated E3 ubiquitin ligase involved in RNA splicing and DNA damage response; reported to be overexpressed in lung cancer and to promote resistance to cisplatin and BCL-2 inhibitor [58]. |  | 1.38 | 1.62 * | 2.23 *** | Monotonically upregulated | 27 | 57 | 75 |
| A-kinase anchor protein 8-like (**AKAP8L**) | Nuclear scaffold protein involved in RNA processing and chromatin organization; promotes cell growth via interaction with the mTORC1 signaling complex and is associated with poor prognosis in various cancers [59]. |  | 1.04 | 1.54 * | 1.61 ** | Monotonically upregulated | 46 | 75 | 93 |
| Probable ATP-dependent RNA helicase DDX59 (**DDX59**) | ATP-dependent RNA helicase involved in RNA metabolism. In lung cancer models, DDX59 has been reported to be induced via EGFR/Ras-MAPK signalling and to contribute to tumor growth [60]. |  | 1.57 | 1.54<br>* | 2.43<br>* | Monotonically upregulated | 24 | 36 | 68 |
| DNA-binding protein RFX5 (**RFX5**) | DNA-binding transcription factor regulating MHC class II genes; reported as highly expressed in MM cell lines, associated with poorer survival in independent patient cohorts, and overexpressed in relapsed/refractory MM [11,61]. |  | 1.56 | 1.52<br>* | 2.37<br>*** | Monotonically upregulated | 27 | 31 | 65 |
| CD320 antigen (**CD320**) | Transcobalamin receptor mediating cobalamin uptake; reported to be overexpressed in MM PCs, associated with poor prognosis, and proposed as a potential CAR T-cell therapeutic target [62]. |  | 1.38 | 2.74<br>* | 3.79<br>*** | Monotonically upregulated | 19 | 21 | 54 |
| Nucleolin (**NCL**) | Multifunctional nucleolar RNA-binding protein regulating RNA metabolism and ribosome biogenesis; reported to promote survival in B-cell lymphoma cells [63] and NF-κB-mediated bortezomib resistance in human MM cell lines [64]. | Anti-nucleolin aptamer - AS1411 (Clinical-stage, Phase II) | 1.09 | 1.70<br>** | 1.85<br>*** | Monotonically upregulated | 37 | 32 | 66 |
| Serine/arginine-rich splicing factor 1 (**SRSF1**) | Serine/arginine-rich splicing factor; reported to be overexpressed in MM and associated with poor prognosis, promoting oncogenic splicing programs that support MM cell survival [65]. |  | 1.12 | 1.61<br>** | 1.80<br>*** | Monotonically upregulated | 44 | 38 | 68 |
| Thymidine kinase, cytosolic (**TK1**) | Cell-cycle-regulated enzyme involved in nucleotide metabolism; elevated serum TK1 levels have been positively correlated with increased proliferative activity and aggressive disease features in MM [66]. | CDK4/6 inhibitor - Palbociclib (FDA approved; indirect modulation of TK1 expression) | 1.24 | 2.42<br>* | 3.00<br>*** | Monotonically upregulated | 0 | 0 | 21 |
| Intercellular adhesion molecule 3 (**ICAM3**) | Type I transmembrane ICAM adhesion molecule mediating immune cell interactions; highly expressed in malignant PCs in MM and proposed as a therapeutic target and liquid biopsy biomarker [22,67]. |  | 1.26 | 1.66<br>* | 2.09<br>*** | Monotonically upregulated | 52 | 23 | 27 |
| Deoxyuridine 5'-triphosphate | Nucleotide metabolism enzyme preventing uracil misincorporation into DNA; reported | dUTPase inhibitor - TAS- | 1.07 | 1.50<br>* | 1.62<br>*** | Monotonically | 67 | 29 | 27 |
| nucleotidohydrolase, mitochondrial<br>**(DUT)** | to promote bortezomib resistance in MM, with its high expression associated with poor prognosis [68]. | 114<br>(Clinical-stage, Phase I) |  |  |  | upregulated |  |  |  |
| DNA topoisomerase 2-beta<br>**(TOP2B)** | DNA topoisomerase resolving DNA topology during replication and transcription; genome-wide CRISPR screening has shown that TOP2B depletion resensitizes drug-resistant MM cells to immunomodulatory agents [69]. | Topoisomerase II inhibitors (e.g., doxorubicin; FDA approved) | 1.17 | 1.51<br>* | 1.77<br>** | Monotonically upregulated | 75 | 25 | 23 |
| Small ubiquitin-related modifier 2<br>**(SUMO2)** | Ubiquitin-like modifier regulating protein function via SUMOylation, affecting DNA repair and stress responses; reported to be upregulated in MM within an activated SUMO pathway associated with relapse and resistance to proteasome inhibitors [70]. | SUMO pathway inhibitors (e.g., subasumstat / TAK-981; Clinical-stage, Phase I/II) | 1.23 | 1.89<br>* | 2.32<br>*** | Monotonically upregulated | 121 | 45 | 47 |
| Peptidoglycan recognition protein 1<br>**(PGLYRP1)** | Secreted innate immune protein mediating cell death and immune activation; reduced in an MM bone marrow coculture model, with decreased levels linked to diminished innate immune responses against MM cells [71]. |  | 0.73 | 0.22<br>* | 0.16<br>*** | Monotonically downregulated | 43 | 50 | 72 |
| Cathelicidin antimicrobial peptide<br>**(CAMP)** | Secreted innate immune protein activating Wnt/β-catenin signalling; promotes osteogenic differentiation and reduces bone resorption [72–74]. | | 0.76 | 0.21<br>* | 0.16<br>*** | Monotonically downregulated | 39 | 61 | 66 |
| Cathepsin G<br>**(CTSG)** | Secreted and membrane-bound serine protease regulating cell death, immune cytotoxicity, and platelet-immune interactions via RAGE and PAR4 signaling [75]. In MM, lower expression has been associated with high-risk disease and adverse prognosis [76]. |  | 0.80 | 0.21<br>** | 0.17<br>*** | Monotonically downregulated | 36 | 56 | 60 |
| Lactotransferrin<br>**(LTF)** | Secreted iron-binding protein regulating iron homeostasis and inflammatory signalling (TLR4/NF-κB); reported to promote osteoblast activity and inhibit osteoclastogenesis [77,78]. | | 0.75 | 0.29<br>* | 0.22<br>*** | Monotonically downregulated | 44 | 54 | 67 |
| Synaptic vesicle membrane protein VAT-1 homolog | Quinone oxidoreductase-family protein that negatively regulates mitochondrial fusion in cooperation with mitofusin proteins |  | 0.82 | 0.58<br>* | 0.47<br>*** | Monotonically downregulated | 62 | 22 | 32 |

| (VAT1) | (MFN1/2). |  |  |  |  |  |  |  |  |
| --- | --- | --- | --- | --- | --- | --- | --- | --- | --- |
| Switch-associated protein 70 (SWAP70) | Regulator of actin assembly driven by the small GTPase RAC1 at membrane remodeling sites, supporting phagocytosis [79]. |  | 0.52 | 0.34 * | 0.18 *** | Monotonically downregulated | 34 | 26 | 0 |

<sup>a</sup> Statistical significance is indicated as follows: \*adjusted p-value < 0.05; \*\*adjusted p-value < 0.01; \*\*\*adjusted p-value < 0.001.
<sup>b</sup> Degree values represent the number of connected neighbours in the stage-specific co-expression networks. SWAP70 and TK1 were retained as isolated nodes (degree set to 0) in stages where they showed low detection frequency: SWAP70 in MM (9/45 samples) and TK1 in MGUS and SMM (1/29 and 3/20 samples, respectively) (**Methods 2.7.3.**)

Investigation of co-expression relationships among the prioritised proteins at the MM stage revealed three distinct components with internal connections between the proteins (**Figure 3**).

**Figure 3:**
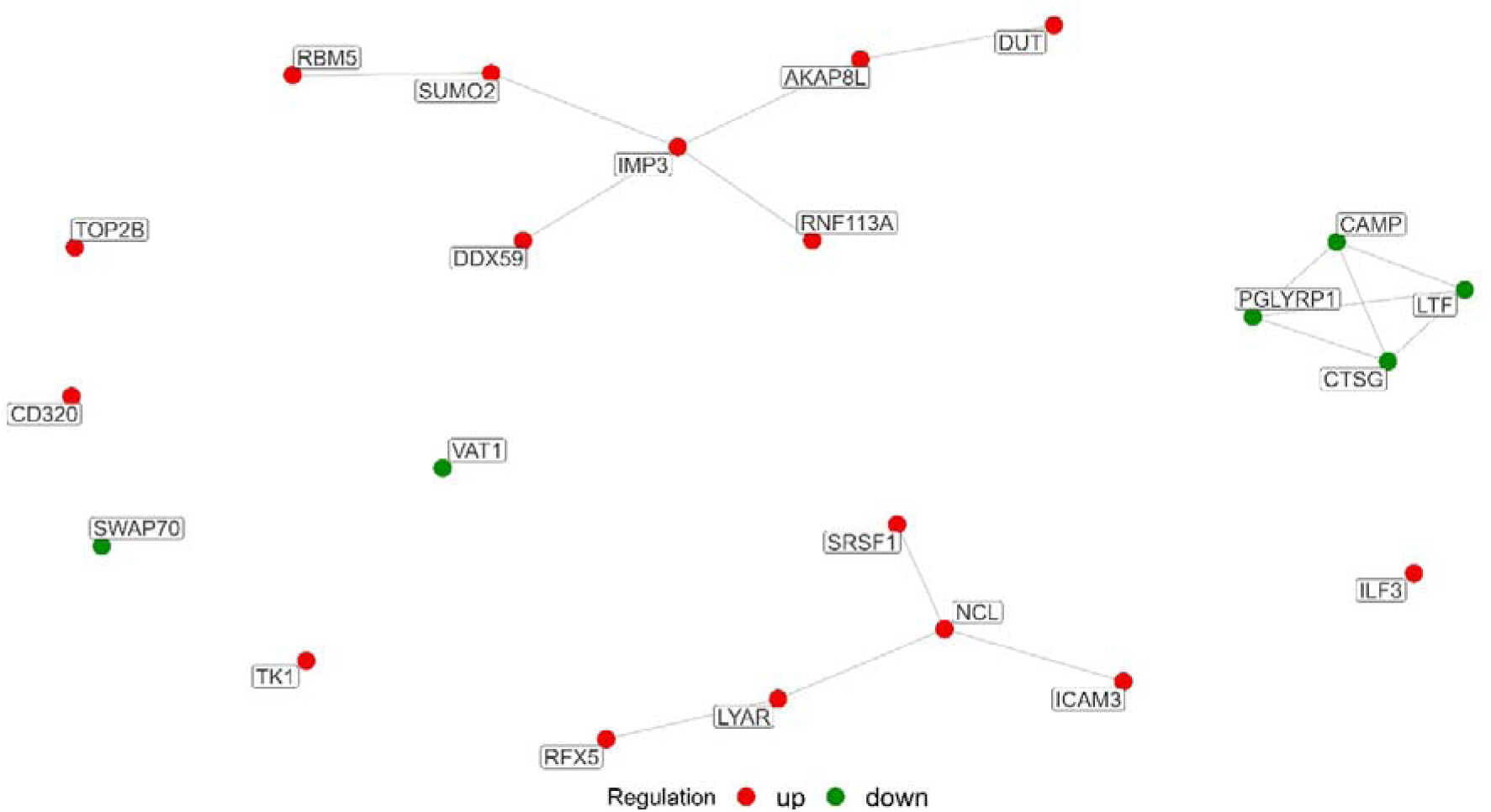
Co-expression subnetwork of the 22 prioritised proteins in MM. Nodes represent the prioritised proteins, and edges denote strong pairwise co-expression relationships amongst them in MM. Node colour indicates regulation status (red = monotonically upregulated; green = monotonically downregulated).

As representative examples, the stage-specific co-expression subnetworks of CTSG, reflecting immune-associated functional programs, nucleolin (NCL) and U3 small nucleolar ribonucleoprotein IMP3 (IMP3), representing nucleolar organisation, ribosome biogenesis and rRNA processing, are further shown in **Figure 4**.

**Figure 4:**
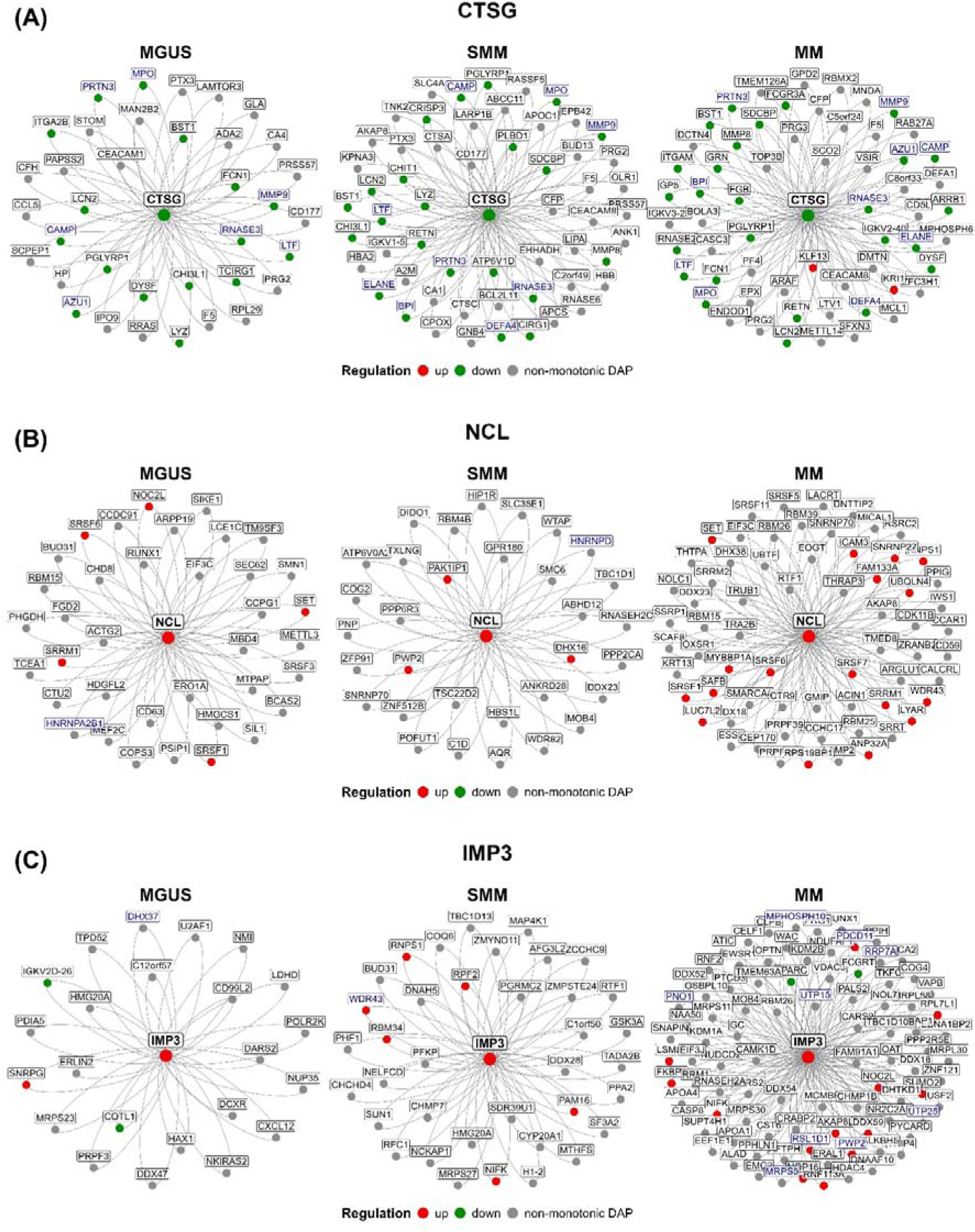
Stage-specific co-expression neighbourhoods of representative prioritised proteins (A) CTSG, (B) NCL, and (C) IMP3. For each protein, first-order co-expression subnetworks are shown across MGUS, SMM, and MM (left to right). The central node represents the protein of interest, and surrounding nodes correspond to its direct co-expression neighbours in the respective stage. Edges denote strong pairwise co-expression relationships. Node colour indicates regulation status (red = monotonically upregulated; green = monotonically downregulated; grey = non-monotonic DAP). Protein labels shown in blue indicate neighbours that also share a direct PPI with the central protein.

CTSG, a secreted and membrane-bound serine protease, was monotonically downregulated across disease stages, yet its connectivity within the co-expression network increased during disease progression, with the largest change occurring between MGUS and SMM (**Figure 4A**). In MGUS, its co-expression neighbours were primarily associated with neutrophil degranulation and coagulation pathways, with part of these relationships also supported by known CTSG protein-protein interaction partners (**Figure 4A**). In SMM and MM, the network further expanded to include proteins linked to the complement cascade, extracellular matrix organisation, and platelet-related pathways. Among the neighbours consistently co-expressed with CTSG across all disease stages were the prioritised secreted proteins LTF, peptidoglycan recognition protein 1 (PGLYRP1), and cathelicidin antimicrobial peptide (CAMP), which also exhibited a monotonic reduction in abundance.

NCL was upregulated across disease stages and showed relatively stable connectivity between MGUS and SMM, followed by a marked expansion of its co-expression neighbourhood in MM (**Figure 4B**). In MGUS and SMM, its co-expression neighbours were primarily RNA-binding and spliceosome-associated proteins involved in mRNA processing and RNA splicing. At the MM stage, NCL became embedded within a broader and denser co-expression network that incorporated additional proteins associated with RNA metabolism, transcriptional regulation, and nucleolar organisation. Among the neighbours observed in the MM network were the prioritised proteins serine/arginine-rich splicing factor 1 (SRSF1), cell growth-regulating nucleolar protein (LYAR), and intercellular adhesion molecule 3 (ICAM3), which likewise increased monotonically in abundance.

IMP3 was monotonically upregulated across disease stages and showed progressive expansion of its co-expression neighbourhood from MGUS to MM (**Figure 4C**). In MGUS, the network was limited and mainly comprised proteins associated with pre-mRNA splicing and transcription-related functions. In SMM, the neighbourhood incorporated additional nucleolar factors involved in early ribosome assembly. At the MM stage, the network expanded substantially and was dominated by proteins involved in rRNA maturation and ribosomal subunit production, also supported by known protein-protein interactions (**Figure 4C**). The network also included proteins associated with translational processes, including components of mitochondrial ribosomal complexes. Among the co-expressed neighbours in MM were the prioritised proteins small ubiquitin-related modifier 2 (SUMO2), A-kinase anchor protein 8-like (AKAP8L), probable ATP-dependent RNA helicase DDX59 (DDX59), and E3 ubiquitin-protein ligase RNF113A (RNF113A), all monotonically increasing across disease stages.

### 3.5. Independent validation of prioritised proteins

#### 3.5.1. Transcriptomic evidence supporting the proteomic findings

To place our prioritised findings (shortlist of 22 proteins) in the context of previously reported molecular alterations during MM development, publicly available transcriptomic data were examined. Following a review of the main text and the corresponding datasets, 3 studies analysing differential expression of CD138⁺ PCs from healthy donors and patients with MGUS, SMM or MM were identified. These included a meta-analysis integrating 12 GEO microarray datasets of CD138⁺ PCs from healthy donors, MGUS and MM patients (GSE1395, GSE47552, GSE5900, GSE6477, GSE61597, GSE13951, GSE2113, GSE24870, GSE27838, GSE6474, GSE39754, GSE7116) [10], an RNA-seq study profiling CD138⁺ PCs from MM patients and healthy donors (GSE153381) [11], and a computational transcriptomic analysis reporting differential expression results from 4 independent GEO microarray datasets of CD138⁺ PCs (GSE5900, GSE6477, GSE13591, GSE47552) [48] **(Additional file 6)**.

From these studies, differential expression results were retrieved for the following comparisons (**Additional file 6**):

- MGUS vs healthy controls (meta-analysis [10]; GSE5900, GSE6477, GSE13591, GSE47552 [48]),
- SMM vs healthy controls (GSE5900, GSE6477, GSE47552 [48]),
- MM vs MGUS (meta-analysis [10]), and
- MM vs healthy controls (meta-analysis [10]; GSE153381 [11]; GSE6477, GSE13591, GSE47552 [48]).

To capture alterations emerging at early stages and persisting during progression to MM, in line with the objective of our study, transcriptomic changes were considered, if detected as differentially expressed in at least one precursor-stage comparison (MGUS vs healthy or SMM vs healthy) and in at least one MM comparison (MM vs healthy or MM vs MGUS), with the same direction of regulation as the proteomic trend.

According to these criteria, 7 of the 22 prioritised proteins (32%) were supported by transcriptomic evidence (RFX5, CD320, DUT, CAMP, CTSG, LTF, and SWAP70), thereby reinforcing the validity of the proteomic findings. Detailed results of the validation analysis are provided in **Additional file 6**.

#### 3.5.2. Validation of CTSG and LTF progressive downregulation across disease stages

In an effort to further experimentally validate the robustness and reproducibility of our -omics observations, we conducted additional expression analyses of selected proteins using complementary methodological approaches and an independent patient cohort. Specifically, we employed MFC to assess the surface expression of CTSG on clonal PCs and ELISA to quantify the levels of secreted LTF in patients’ BM plasma.

CTSG exhibited a consistently low but detectable level of expression across BM nucleated cells, without a distinct separation between positive and negative populations within individual cell subsets. Due to this lack of a clear bimodal distribution, we evaluated the median fluorescence intensity (MFI) of CTSG on clonal PCs and compared it between the three stages. To ensure internal normalization and account for inter-sample variability, MFI values were compared against those measured in erythroblasts, which served as an internal baseline/negative control in each sample (**Additional file 7**) [80]. Our analysis revealed a progressive decrease in CTSG expression in clonal PCs along the disease continuum, from asymptomatic conditions to overt MM, approaching statistical significance (p-value = 0.0517; **Figure 5A**). In the same line, the ELISA-based measurements demonstrated that LTF levels were statistically significantly reduced in the BM plasma of NDMM patients compared with MGUS (p-value = 0.0106) and showed a similar decreasing trend compared with SMM, narrowly missing statistical significance (p-value = 0.063; **Figure 5B**), supporting the proteomics findings of LTF systematic stage-dependent downregulation during disease progression.

**Figure 5:**
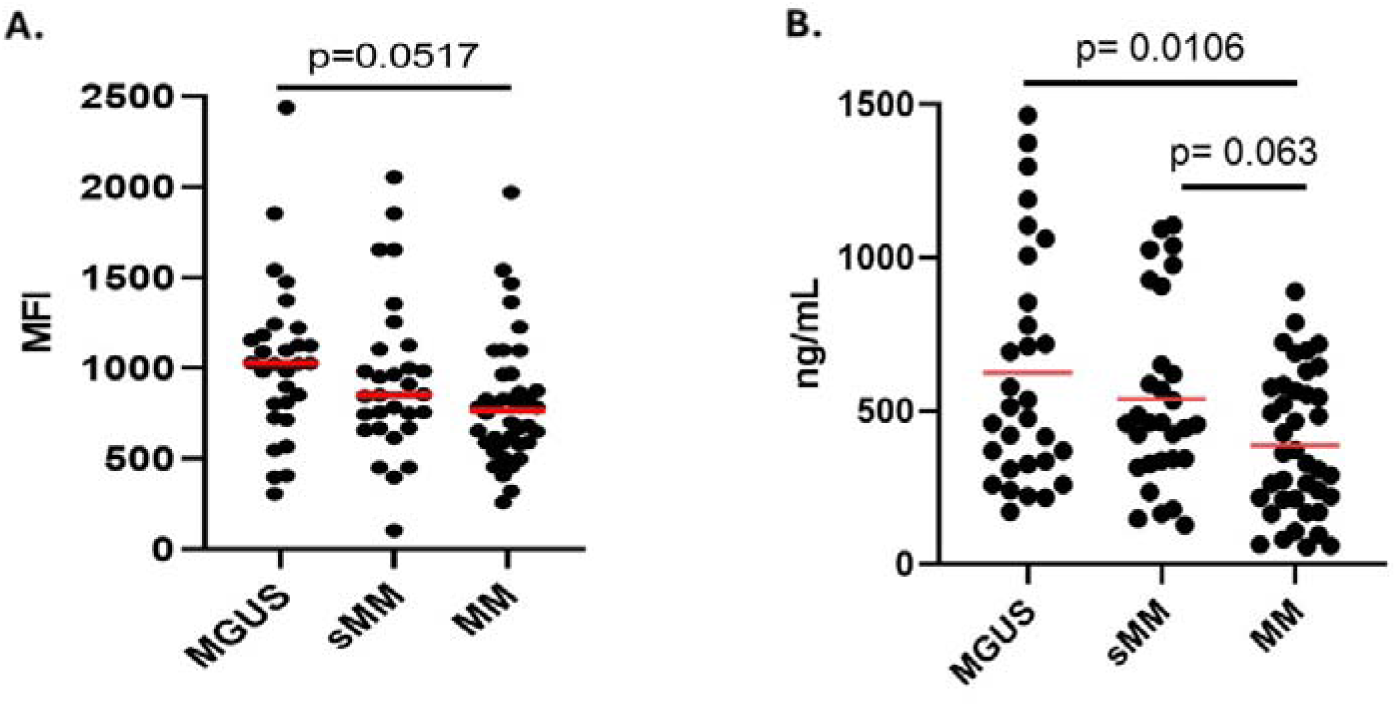
Independent validation of CTSG and LTF across disease stages. **(A)** Surface expression of CTSG on BM clonal PCs and (**B**) levels of LTF in BM plasma of patients with MGUS (n = 30), SMM (n = 30) and MM (n = 40). Red horizontal lines show mean values. Statistical significance was tested with one-way ANOVA followed by Tukey’s test for pairwise comparisons. MFI: median fluorescence intensity.

## 4. DISCUSSION

Despite major advances in understanding MM biology, the molecular mechanisms underlying the transition from precursor conditions to symptomatic disease remain incompletely characterised. While genomic and transcriptomic studies have provided important insights into disease evolution, relatively few have examined these processes at protein level [20]. In this study, we investigated proteomic alterations across the MGUS-SMM-MM spectrum in BM-derived CD138⁺ PCs using differential abundance and network-based analyses. This approach identified 370 proteins exhibiting monotonic changes across the three disease stages, with abundance progressively increasing or decreasing during disease progression (**Figure 1**).

Pathway and protein-protein interaction analyses (**Figure 2**) revealed that monotonically upregulated proteins were mainly associated with pathways related to celllllcycle progression, ribosome biogenesis, MYC target gene sets, and the UPR. These processes are tightly linked to the biology of malignant PCs, which require high transcriptional and translational output to sustain uncontrolled proliferation and the production of large quantities of monoclonal immunoglobulin. The enrichment of ribosome biogenesis and MYClllassociated pathways is consistent with MYC’s established role as a central oncogenic driver in MM, promoting cell growth, anabolic metabolism, and protein translation to support the biosynthetic demands of malignant PCs [81]. At the same time, activation of the UPR reflects the increased burden on the endoplasmic reticulum (ER) and aligns with the known sensitivity of myeloma cells to proteasome inhibitors [82]. In contrast, monotonically downregulated proteins were primarily associated with immunelllrelated pathways, suggesting a progressive impairment of immune signalling and functional reprogramming during the transition from MGUS to clinically active disease. This pattern is compatible with the ability of malignant PCs to evade immune surveillance by disrupting interactions with key immune components such as T cells and dendritic cells, through mechanisms such as the downregulation of the antigen processing machinery [83,84].

To further characterise the behaviour of proteins exhibiting monotonic abundance changes during disease progression, co-expression network analysis was performed to identify proteins whose abundance levels change simultaneously in a coordinated manner (either in the same or opposite direction) across the three disease stages. Based on a combination of criteria including co-expression patterns, detection frequency, biological relevance, and translational potential, 22 representative proteins were prioritised (**Table 2**). These proteins included established MM-associated factors alongside additional candidates with potential relevance for biomarker discovery, therapeutic targeting, and mechanistic insights into MM.

Among the prioritised candidates, four proteins (CTSG, LTF, CAMP, and PGLYRP1) involved in innate immune responses were identified, all showing monotonic downregulation across disease stages and strong co-expression within the MM subnetwork of prioritised proteins (**Figure 3**). Of note, lower levels of CTSG have also been associated with high-risk MM in bioinformatic risk models [76]. A closer examination of its stage-specific co-expression neighbourhoods (**Figure 4A**) showed that, despite its decreasing abundance, CTSG exhibited increasing connectivity across disease stages and became progressively co-expressed with proteins involved in immune and microenvironment-related pathways. This pattern suggests that reduced CTSG levels and potentially CTSG activity may contribute to immune and microenvironmental dysregulation during disease progression. Consistent with the known roles of CTSG in innate immune but also cytotoxic responses [75], such dysregulation may thus involve altered complement and inflammatory signalling together with diminished cytotoxic and cell death responses, potentially favouring malignant PC survival during MM progression.

To assess the clinical relevance of these observations, the downregulation of CTSG and one of its co-expression neighbours, LTF, across disease stages was validated in independent clinical samples by flow cytometry and ELISA, respectively, as suitable antibodies and assays were commercially available. LTF is an iron-binding glycoprotein of the transferrin family involved in bone homeostasis [77,78] and inflammatory responses, in which it has been reported to act synergistically with CTSG [85]. Both CTSG and LTF showed a gradual decrease from MGUS and SMM to MM, which was further supported by transcriptomic data [10,11,48] (**Additional file 6**), highlighting their potential as candidate biomarkers of disease progression, meriting further investigation.

Another prominent candidate among the prioritised proteins was NCL, a nucleolar RNA-binding protein that was upregulated across disease stages. In our dataset, NCL exhibited a marked expansion of its co-expression neighbourhood and showed increased co-expression with RNA-binding and spliceosome-associated proteins in MM compared with earlier stages (**Figure 4B**). This is consistent with the established role of NCL in RNA metabolism, including the regulation of pre-mRNA processing and RNA maturation [86], processes central to gene expression control in highly secretory malignant PCs which sustain high levels of monoclonal immunoglobulin production [87]. Notably, previous studies have shown that NCL promotes survival in B-cell lymphoma cells [63], while its inhibition reduces proliferation and increases apoptosis in MM cell lines [64]. Moreover, the co-expression neighbourhood in MM includes prioritised proteins implicated in MM biology, such as the oncogenic factors SRSF1 [65] and LYAR [57], and the adhesion molecule ICAM3, highly expressed in MM cells [22]. Collectively, these findings support a potential role for NCL in RNA regulatory processes associated with malignant PC survival and MM progression. NCL also represents an emerging pharmacologically tractable target, as the nucleolin-targeting aptamer AS1411 has been investigated in a Phase II clinical trial in acute myeloid leukemia (ClinicalTrials.gov identifier: NCT00512083), while more recent evidence demonstrates anti-tumor activity of AS1411-based NCL-targeting drug conjugates in esophageal cancer cell lines and mouse xenograft models [88].

An additional prioritised protein of interest in our study was IMP3, a component of the U3 small nucleolar ribonucleoprotein complex involved in early pre-rRNA processing during ribosome biogenesis [89]. To our knowledge, its role in MM progression has not previously been investigated. Along with its monotonic increase in abundance across disease stages, IMP3 displayed a progressive expansion of its co-expression neighbourhood (**Figure 4C**), increasingly enriched for proteins involved in rRNA maturation, ribosomal subunit production, and translational processes. These observations support a progressive involvement of IMP3 in the nucleolar ribosome biogenesis machinery during MM progression, a process closely linked to MYC-driven growth in malignant PCs [81]. Within the IMP3 co-expression neighbourhood in MM, prioritised proteins previously implicated in malignancy-associated mechanisms were also identified. Among them, SUMO2 has been linked to activation of the SUMO pathway, which has emerged as a potential therapeutic target in MM [70], while AKAP8L has been associated with mTORC1-driven cell growth signalling [59]. Additional neighbours include DDX59, reported to promote tumour growth through Ras–MAPK signalling [60], and RNF113A, a spliceosome-associated ubiquitin ligase overexpressed in several cancers [58]. In this context, IMP3 may represent an underexplored factor supporting malignant PC growth.

Two additional prioritised candidates, the synaptic vesicle membrane protein VAT-1 homolog (VAT1) and the switch-associated protein 70 (SWAP70), were identified as proteins of potential interest that warrant further investigation. Both displayed monotonic downregulation across disease stages accompanied by reduced co-expression network connectivity, suggesting more independent changes in protein abundance during disease progression. VAT1 has been reported to negatively regulate mitochondrial fusion through interactions with mitofusin proteins [90] and has also been suggested to participate in phospholipid transfer between the ER and mitochondrial membranes [91,92]. Its decreased abundance across disease stages may suggest increased mitochondrial fusion and alterations in the mitochondrial membrane phospholipid composition during MM progression, potentially influencing mitochondrial function and metabolic adaptation in malignant PCs. In parallel, SWAP70 is a RAC1-responsive regulator of actin cytoskeleton remodelling through interactions with phosphoinositides and F-actin. Its reduced abundance during disease progression may therefore be associated with changes in cytoskeletal organisation and RAC1-dependent signalling, processes known to influence cell adhesion and interactions within the BM microenvironment [79].

Collectively, this study provides important insights into proteomic alterations accompanying progression from MGUS and SMM to symptomatic MM, although certain limitations should be acknowledged. The cohort size was moderate, and the cross-sectional study design does not allow direct monitoring of disease progression within the same patients over time. Despite these limitations, the agreement between the proteomic findings and independent transcriptomic datasets, together with the validation of selected candidates, supports the biological relevance of the results. To our knowledge, this is the first proteomic study in BM-derived CD138⁺ PCs directly comparing the MGUS, SMM, and MM disease stages. Overall, our results highlight coordinated changes in immune regulation, RNA processing, and ribosome biogenesis as central features of MM progression and identify candidate proteins, including CTSG, LTF, NCL and IMP3, in parallel with their predicted network partners, thus opening the way towards new avenues for mechanistic investigation, biomarker development and therapeutic targeting in MM.

## 5. CONCLUSIONS

The proteomic analysis performed in this study, comparing BM-derived CD138⁺ PCs across MGUS, SMM and MM, provides new insights into the molecular events underlying disease progression. Our findings highlight coordinated molecular alterations associated with MM evolution and support the potential relevance of candidate proteins, including CTSG, LTF, NCL, and IMP3. Supported by independent datasets and experimental validation, these findings can serve as a basis for future studies aimed at elucidating the molecular mechanisms underlying MM progression.

## Supporting information

Addiitonal File 2

## Data Availability

The data supporting the conclusions of this article are included within the article and its additional files. The mass spectrometry proteomics data have been deposited to the ProteomeXchange Consortium via the PRIDE partner repository with the dataset identifier PXD078279.

## 6. LIST OF ABBREVIATIONS

ARI: Adjusted Rand Index
BH: Benjamini–Hochberg
BM: Bone marrow
CAA: 2-chloroacetamide
DAP: Differentially abundant protein
DIA: Data-independent acquisition
dia-PASEF: Data-independent acquisition with parallel accumulation - serial fragmentation
ELISA: Enzyme-Linked Immunosorbent Assay
ER: Endoplasmic Reticulum
FC: Fold Change
FDR: False discovery rate
GO-BP: Gene Ontology Biological Process
IMWG: International Myeloma Working Group
LC-MS/MS: Liquid chromatography - tandem mass spectrometry
MFC: Multi-color flow cytometry
MFI: Median fluorescence intensity
MGUS: Monoclonal gammopathy of undetermined significance
MM: Multiple myeloma
MSigDB: Molecular Signatures Database
MS: Mass spectrometry
NDMM: Newly diagnosed multiple myeloma
PCA: Principal Component Analysis
PCs: Plasma cells
PPI: Protein-protein interaction
ppm: Parts per million
SMM: Smouldering multiple myeloma
SPEED: Sample Preparation by Easy Extraction and Digestion
TCEP·HCl: Tris(2-carboxyethyl)phosphine hydrochloride
TFA: Trifluoroacetic acid
UPR: Unfolded Protein Response

## 7. ADDITIONAL FILES

### Additional file 1

- **File format:** .xlsx
- **Title:** Processed proteomics dataset and differential abundance statistics across MGUS, SMM, and MM.
- **Description:** Processed proteomics data after normalisation, filtering, imputation, and log_2_ transformation, together with pairwise differential abundance results for all pairwise comparisons across stages. Sheet 1 contains the full processed dataset, while Sheet 2 contains the same information restricted to the 370 monotonic DAPs.

### Additional file 2

- **File format:** .xlsx
- **Title:** Pathway enrichment analysis of monotonic DAPs.
- **Description:** Results of pathway enrichment analysis performed separately for monotonically upregulated and downregulated DAPs using GO Biological Process, Reactome, and Hallmark gene sets.

### Additional file 3

- **File format:** .xlsx
- **Title:** Protein–protein interaction network data for the proteomics dataset.
- **Description:** Protein–protein interaction (PPI) network information for proteins in the processed proteomics dataset. For each protein, the degree and corresponding interacting partners are reported.

### Additional file 4

- **File format:** .xlsx
- **Title:** Stage-specific co-expression network data for monotonic DAPs.
- **Description:** Co-expression network information for monotonic DAPs across disease stages. Sheet 1 summarizes connectivity metrics and degree changes. Sheets 2–4 report stage-specific protein degrees and corresponding neighbours.

### Additional file 5

- **File format:** .tiff
- **Title:** Co-expression network visualisation of monotonic DAPs with major connectivity changes across MGUS and MM.
- **Description:** Co-expression subnetworks of monotonic DAPs with the largest positive (Δ > 0) and negative (Δ < 0) connectivity changes between MGUS and MM. Networks are shown separately for MGUS and MM and include the top connectivity-changing monotonic DAPs and their first-order neighbours.

### Additional file 6

- **File format:** .xlsx
- **Title:** Transcriptomic validation data for prioritised monotonic network-rewired DAPs.
- **Description:** Information on the publicly available transcriptomic datasets used for validation and the assessment of transcriptomic support for the 22 prioritised monotonic network-rewired DAPs. Sheet 1 summarises the included studies and datasets, while Sheet 2 reports the differential expression evidence across the selected comparisons.

### Additional file 7

- **File format:** .pdf
- **Title:** Representative flow cytometry histograms of CTSG expression in clonal plasma cells.
- **Description:** Histograms showing the expression levels of CTSG on BM clonal PCs from representative cases of MGUS (blue), SMM (green) and MM (orange) patients. The expression levels of CTSG on erythroblasts (red) in each BM sample was used as an internal baseline/negative control. The relevant MFI values (Median) of CTSG are shown in the Table.

## 8. DECLARATIONS

### 8.1. Ethics approval and consent to participate

All patients provided written informed consent for biological material collection, clinical data collection and analysis, after approval by the institution’s Ethics/Scientific Committee of Alexandra Hospital (No 111368/25.10.2022). The study was conducted in accordance with the ethical principles of the Declaration of Helsinki.

### 8.2. Consent for publication

Not applicable.

### 8.3. Availability of data and materials

The data supporting the conclusions of this article are included within the article and its additional files. The mass spectrometry proteomics data have been deposited to the ProteomeXchange Consortium via the PRIDE [93] partner repository with the dataset identifier PXD078279.

### 8.4. Competing Interests

The authors declare that they have no competing interests.

### 8.5. Funding

This project was supported by the HORIZON-MISS-2021-Cancer Grant #101097094 (ELMUMY).

### 8.6. Authors’ contributions

**Conceptualization**, A.V., J.Z. and E.K.; **Methodology**, F.P., C.G., J.C. and I.K.; **Formal analysis**, F.P., C.G., T.M. and E.G.; **Investigation**, F.P., J.C., P.K., P.B., A.M. and C.L.; **Resources**, E.K., C.L. and O.T.; **Data curation**, F.P., A.T., C.G., J.C. and P.K.; **Writing – original draft preparation**, F.P., A.T. and C.G.; **Writing – review & editing**, F.P., A.T., C.G., T.M., E.G., P.B., I.K., J.C., P.K., O.T., A.V., J.Z. and E.K.; **Visualisation**, F.P., C.G., E.G. and P.B.; **Supervision**, A.V., C.S., J.M. and J.Z.; **Project administration**, A.V. and J.Z.; **Funding acquisition**, A.V., J.Z., O.T., M.D., E.K.

## 8.7 Acknowledgements

Not applicable.

---

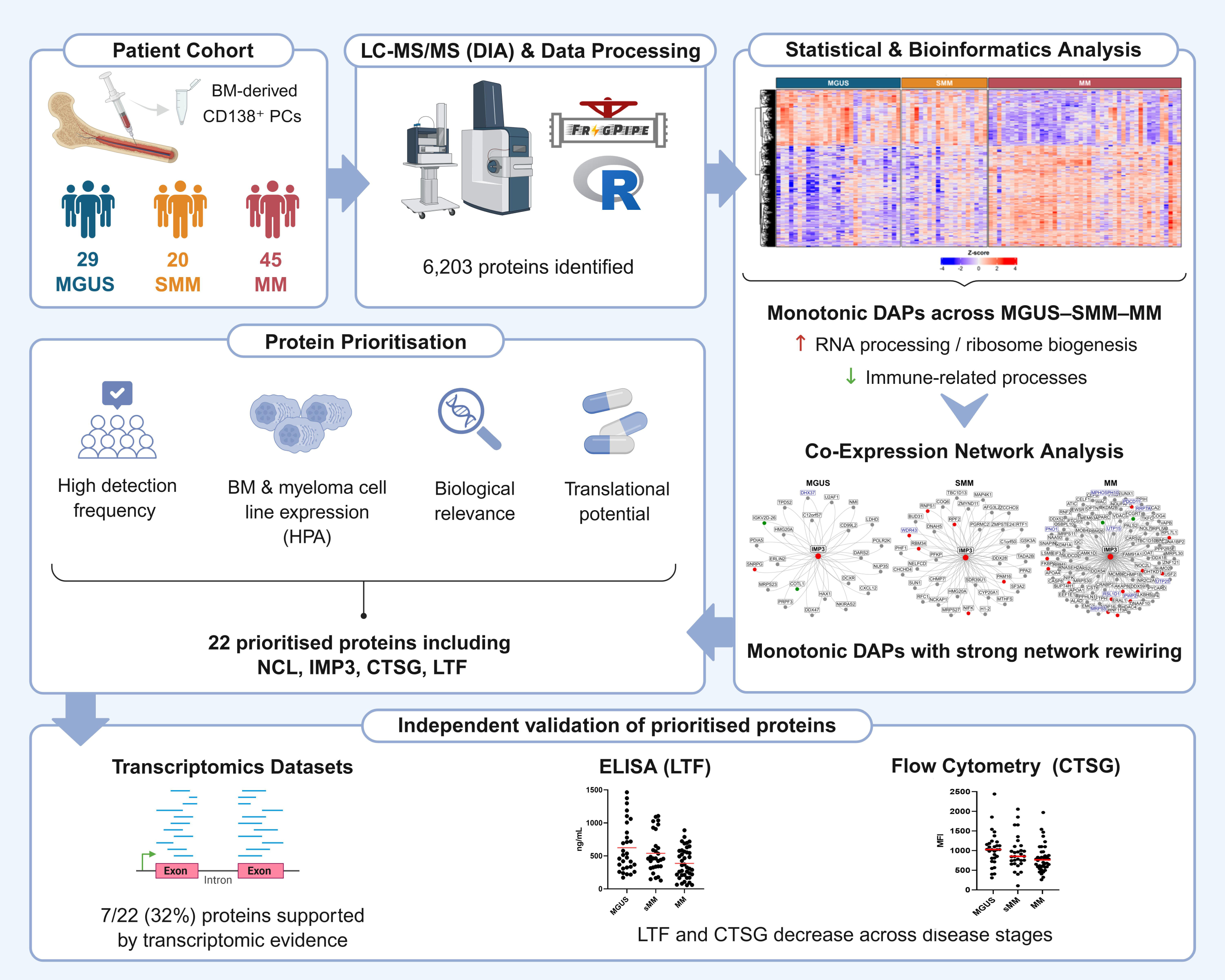

## Notes

### Competing Interest Statement

The authors have declared no competing interest.

### Author Declarations

The Ethics/Scientific Committee of Alexandra Hospital (No. 111368/25.10.2022) gave ethical approval for this work. The study was conducted in accordance with the Declaration of Helsinki.

## REFERENCES

[1] Rajkumar SV, Dimopoulos MA, Palumbo A, Blade J, Merlini G, Mateos M-V, et al. International Myeloma Working Group updated criteria for the diagnosis of multiple myeloma. Lancet Oncol 2014;15:e538–48. 10.1016/S1470-2045(14)70442-5.

[2] Rajkumar SV. Multiple myeloma: 2022 update on diagnosis, risk stratification, and management. Am J Hematol 2022;97:1086–107. 10.1002/ajh.26590.

[3] Padala SA, Barsouk A, Barsouk A, Rawla P, Vakiti A, Kolhe R, et al. Epidemiology, Staging, and Management of Multiple Myeloma. Medical Sciences 2021;9:3. 10.3390/medsci9010003.

[4] Mateos M-V, Landgren O. MGUS and Smoldering Multiple Myeloma: Diagnosis and Epidemiology. Cancer Treat Res 2016;169:3–12. 10.1007/978-3-319-40320-5_1.

[5] Ho M, Bianchi G, Anderson KC. Proteomics-inspired precision medicine for treating and understanding multiple myeloma. Expert Rev Precis Med Drug Dev 2020;5:67–85. 10.1080/23808993.2020.1732205.

[6] Pinto V, Bergantim R, Caires HR, Seca H, Guimarães JE, Vasconcelos MH. Multiple Myeloma: Available Therapies and Causes of Drug Resistance. Cancers (Basel) 2020;12:407. 10.3390/cancers12020407.

[7] Gong L, Qiu L, Hao M. Novel Insights into the Initiation, Evolution, and Progression of Multiple Myeloma by Multi-Omics Investigation. Cancers (Basel) 2024;16:498. 10.3390/cancers16030498.

[8] Chen M, Jiang J, Hou J. Single-cell technologies in multiple myeloma: new insights into disease pathogenesis and translational implications. Biomark Res 2023;11:55. 10.1186/s40364-023-00502-8.

[9] Maura F, Bergsagel PL, Ziccheddu B, Kumar S, Maclachlan K, Derkach A, et al. Genomics Define Malignant Transformation in Myeloma Precursor Conditions. J Clin Oncol 2026;44:188–99. 10.1200/JCO-25-01733.

[10] Aljabban J, Syed S, Syed S, Rohr M, Mukhtar M, Aljabban H, et al. Characterization of monoclonal gammopathy of undetermined significance progression to multiple myeloma through meta-analysis of GEO data. Heliyon 2023;9:e17298. 10.1016/j.heliyon.2023.e17298.

[11] Alvarez-Benayas J, Trasanidis N, Katsarou A, Ponnusamy K, Chaidos A, May PC, et al. Chromatin-based, in cis and in trans regulatory rewiring underpins distinct oncogenic transcriptomes in multiple myeloma. Nat Commun 2021;12:5450. 10.1038/s41467-021-25704-2.

[12] Zhan F, Barlogie B, Arzoumanian V, Huang Y, Williams DR, Hollmig K, et al. Gene-expression signature of benign monoclonal gammopathy evident in multiple myeloma is linked to good prognosis. Blood 2007;109:1692–700. 10.1182/blood-2006-07-037077.

[13] Chng WJ, Kumar S, Vanwier S, Ahmann G, Price-Troska T, Henderson K, et al. Molecular dissection of hyperdiploid multiple myeloma by gene expression profiling. Cancer Res 2007;67:2982–9. 10.1158/0008-5472.CAN-06-4046.

[14] Agnelli L, Mosca L, Fabris S, Lionetti M, Andronache A, Kwee I, et al. A SNP microarray and FISH-based procedure to detect allelic imbalances in multiple myeloma: an integrated genomics approach reveals a wide gene dosage effect. Genes Chromosomes Cancer 2009;48:603–14. 10.1002/gcc.20668.

[15] López-Corral L, Corchete LA, Sarasquete ME, Mateos MV, García-Sanz R, Fermiñán E, et al. Transcriptome analysis reveals molecular profiles associated with evolving steps of monoclonal gammopathies. Haematologica 2014;99:1365–72. 10.3324/haematol.2013.087809.

[16] Ledergor G, Weiner A, Zada M, Wang S-Y, Cohen YC, Gatt ME, et al. Single cell dissection of plasma cell heterogeneity in symptomatic and asymptomatic myeloma. Nat Med 2018;24:1867–76. 10.1038/s41591-018-0269-2.

[17] Moreno Rueda LY, Wang H, Akagi K, Dang M, Vora A, Qin L, et al. Single-cell analysis of neoplastic plasma cells identifies myeloma pathobiology mediators and potential targets. Cell Rep Med 2025;6:101925. 10.1016/j.xcrm.2024.101925.

[18] Boiarsky R, Haradhvala NJ, Alberge J-B, Sklavenitis-Pistofidis R, Mouhieddine TH, Zavidij O, et al. Single cell characterization of myeloma and its precursor conditions reveals transcriptional signatures of early tumorigenesis. Nat Commun 2022;13:7040. 10.1038/s41467-022-33944-z.

[19] Dang M, Wang R, Lee HC, Patel KK, Becnel MR, Han G, et al. Single cell clonotypic and transcriptional evolution of multiple myeloma precursor disease. Cancer Cell 2023;41:1032–1047.e4. 10.1016/j.ccell.2023.05.007.

[20] Ismail NH, Mussa A, Al-Khreisat MJ, Mohamed Yusoff S, Husin A, Johan MF. Proteomic Alteration in the Progression of Multiple Myeloma: A Comprehensive Review. Diagnostics 2023;13:2328. 10.3390/diagnostics13142328.

[21] Kamal M, Shishido SN, Mason J, Patel K, Manasanch EE, Orlowski RZ, et al. Single-cell proteomic analysis reveals Multiple Myeloma heterogeneity and the dynamics of the tumor immune microenvironment in precursor and advanced states. Neoplasia 2025;66:101189. 10.1016/j.neo.2025.101189.

[22] Setayesh SM, Ndacayisaba LJ, Rappard KE, Hennes V, Rueda LYM, Tang G, et al. Targeted single-cell proteomic analysis identifies new liquid biopsy biomarkers associated with multiple myeloma. NPJ Precis Oncol 2023;7:95. 10.1038/s41698-023-00446-0.

[23] Glavey S V, Naba A, Manier S, Clauser K, Tahri S, Park J, et al. Proteomic characterization of human multiple myeloma bone marrow extracellular matrix. Leukemia 2017;31:2426–34. 10.1038/leu.2017.102.

[24] Janker L, Mayer RL, Bileck A, Kreutz D, Mader JC, Utpatel K, et al. Metabolic, Anti-apoptotic and Immune Evasion Strategies of Primary Human Myeloma Cells Indicate Adaptations to Hypoxia. Mol Cell Proteomics 2019;18:936–53. 10.1074/mcp.RA119.001390.

[25] Ho M, Dasari S, Visram A, Drake MT, Charlesworth MC, Johnson KL, et al. An atlas of the bone marrow bone proteome in patients with dysproteinemias. Blood Cancer J 2023;13:63. 10.1038/s41408-023-00840-8.

[26] Li J, Smith LS, Zhu H-J. Data-independent acquisition (DIA): An emerging proteomics technology for analysis of drug-metabolizing enzymes and transporters. Drug Discov Today Technol 2021;39:49–56. 10.1016/j.ddtec.2021.06.006.

[27] Karousi P, Voumvouraki M, Nikolaou PE, Kollias I, Paradeisi F, Sampanai E, et al. Easy Proteomics Sample Preparation: Technical Repeatability and Workflow Optimization Across 8 Biological Matrices in a New Core Facility Setting. Proteomics 2025;25:15–24. 10.1002/pmic.70064.

[28] Yu F, Teo GC, Kong AT, Fröhlich K, Li GX, Demichev V, et al. Analysis of DIA proteomics data using MSFragger-DIA and FragPipe computational platform. Nat Commun 2023;14:4154. 10.1038/s41467-023-39869-5.

[29] Demichev V, Messner CB, Vernardis SI, Lilley KS, Ralser M. DIA-NN: neural networks and interference correction enable deep proteome coverage in high throughput. Nat Methods 2020;17:41–4. 10.1038/s41592-019-0638-x.

[30] Li K, Teo GC, Yang KL, Yu F, Nesvizhskii AI. diaTracer enables spectrum-centric analysis of diaPASEF proteomics data. Nat Commun 2025;16:95. 10.1038/s41467-024-55448-8.

[31] Bateman A, Martin M-J, Orchard S, Magrane M, Adesina A, Ahmad S, et al. UniProt: the Universal Protein Knowledgebase in 2025. Nucleic Acids Res 2025;53:D609–17. 10.1093/nar/gkae1010.

[32] R Core Team. R: A language and environment for statistical computing. R Foundation for Statistical Computing, Vienna, Austria; 2024. https://www.R-project.org/.

[33] Benjamini Y, Hochberg Y. Controlling the False Discovery Rate: A Practical and Powerful Approach to Multiple Testing. J R Stat Soc Series B Stat Methodol 1995;57:289–300. 10.1111/j.2517-6161.1995.tb02031.x.

[34] Yu G, He Q-Y. ReactomePA: an R/Bioconductor package for reactome pathway analysis and visualization. Mol Biosyst 2016;12:477–9. 10.1039/c5mb00663e.

[35] Yu G, Wang L-G, Han Y, He Q-Y. clusterProfiler: an R package for comparing biological themes among gene clusters. OMICS 2012;16:284–7. 10.1089/omi.2011.0118.

[36] Subramanian A, Tamayo P, Mootha VK, Mukherjee S, Ebert BL, Gillette MA, et al. Gene set enrichment analysis: A knowledge-based approach for interpreting genome-wide expression profiles. Proceedings of the National Academy of Sciences 2005;102:15545–50. 10.1073/pnas.0506580102.

[37] Szklarczyk D, Kirsch R, Koutrouli M, Nastou K, Mehryary F, Hachilif R, et al. The STRING database in 2023: protein–protein association networks and functional enrichment analyses for any sequenced genome of interest. Nucleic Acids Res 2023;51:D638–46. 10.1093/nar/gkac1000.

[38] Shannon P, Markiel A, Ozier O, Baliga NS, Wang JT, Ramage D, et al. Cytoscape: A Software Environment for Integrated Models of Biomolecular Interaction Networks. Genome Res 2003;13:2498–504. 10.1101/gr.1239303.

[39] Safari-Alighiarloo N, Taghizadeh M, Rezaei-Tavirani M, Goliaei B, Peyvandi AA. Protein-protein interaction networks (PPI) and complex diseases. Gastroenterol Hepatol Bed Bench 2014;7:17–31.

[40] Csárdi G, Nepusz T, Müller K, Horvát S, Traag V, Zanini F, et al. igraph for R: R interface of the igraph library for graph theory and network analysis 2025. 10.5281/zenodo.7682609.

[41] Scrucca L, Fraley C, Murphy TB, Adrian E. R. Model-Based Clustering, Classification, and Density Estimation Using mclust in R. Boca Raton: Chapman and Hall/CRC; 2023. 10.1201/9781003277965.

[42] Milacic M, Beavers D, Conley P, Gong C, Gillespie M, Griss J, et al. The Reactome Pathway Knowledgebase 2024. Nucleic Acids Res 2024;52:D672–8. 10.1093/nar/gkad1025.

[43] van Dam S, Võsa U, van der Graaf A, Franke L, de Magalhães JP. Gene co-expression analysis for functional classification and gene–disease predictions. Brief Bioinform 2017:bbw139. 10.1093/bib/bbw139.

[44] Huynh-Thu VA, Irrthum A, Wehenkel L, Geurts P. Inferring Regulatory Networks from Expression Data Using Tree-Based Methods. PLoS One 2010;5:e12776. 10.1371/journal.pone.0012776.

[45] National Library of Medicine. PubMed 2026. https://pubmed.ncbi.nlm.nih.gov/ (accessed January 20, 2026).

[46] EMBL-EBI. ArrayExpress 2026. https://www.ebi.ac.uk/biostudies/arrayexpress (accessed January 20, 2026).

[47] National Center for Biotechnology Information. Gene Expression Omnibus (GEO) 2026. https://www.ncbi.nlm.nih.gov/geo/ (accessed January 20, 2026).

[48] Savva K, Bourdakou MM, Stellas D, Zoidakis J, Spyrou GM. Computational Drug Repurposing Across the Multiple Myeloma Spectrum: From MGUS to MM. Cancers (Basel) 2025;17. 10.3390/cancers17183045.

[49] Terpos E, Kostopoulos I V, Kastritis E, Ntanasis-Stathopoulos I, Migkou M, Rousakis P, et al. Impact of Minimal Residual Disease Detection by Next-Generation Flow Cytometry in Multiple Myeloma Patients with Sustained Complete Remission after Frontline Therapy. Hemasphere 2019;3:e300. 10.1097/HS9.0000000000000300.

[50] Mateos M-V, Kumar S, Dimopoulos MA, González-Calle V, Kastritis E, Hajek R, et al. International Myeloma Working Group risk stratification model for smoldering multiple myeloma (SMM). Blood Cancer J 2020;10:102. 10.1038/s41408-020-00366-3.

[51] Kastritis E, Solia I, Malandrakis P, Theodorakakou F, Ntanasis-Stathopoulos I, Kanellias N, et al. Patterns of progression among 427 patients with smoldering myeloma diagnosed after 2014: importance of monitoring. Blood Adv 2025;9:4444–7. 10.1182/bloodadvances.2025016083.

[52] Human Protein Atlas n.d. https://www.proteinatlas.org (accessed January 20, 2026).

[53] Uhlén M, Fagerberg L, Hallström BM, Lindskog C, Oksvold P, Mardinoglu A, et al. Proteomics. Tissue-based map of the human proteome. Science 2015;347:1260419. 10.1126/science.1260419.

[54] Knox C, Wilson M, Klinger CM, Franklin M, Oler E, Wilson A, et al. DrugBank 6.0: the DrugBank Knowledgebase for 2024. Nucleic Acids Res 2024;52:D1265–75. 10.1093/nar/gkad976.

[55] Jiang F, Tang X, Tang C, Hua Z, Ke M, Wang C, et al. HNRNPA2B1 promotes multiple myeloma progression by increasing AKT3 expression via m6A-dependent stabilization of ILF3 mRNA. J Hematol Oncol 2021;14:54. 10.1186/s13045-021-01066-6.

[56] Zhang Y, Chen X, Xiao Y, Mei Y, Yang T, Li D, et al. Elucidating the role of RBM5 in osteoclastogenesis: a novel potential therapeutic target for osteoporosis. BMC Musculoskelet Disord 2023;24:921. 10.1186/s12891-023-07002-8.

[57] Okamoto H, Mizutani S, Tsukamoto T, Katsuragawa-Taminishi Y, Isa R, Mizuhara K, et al. TAS0612, a Triple Inhibitor for RSK, AKT, and S6K, Is a Promising Anti-Myeloma Agent Regardless of Types of Cytogenetic/Genetic Abnormality. Blood 2023;142:3313–3313. 10.1182/blood-2023-174157.

[58] Shostak K, Jiang Z, Charloteaux B, Mayer A, Habraken Y, Tharun L, et al. The X-linked trichothiodystrophy-causing gene RNF113A links the spliceosome to cell survival upon DNA damage. Nat Commun 2020;11:1270. 10.1038/s41467-020-15003-7.

[59] Zhou L, Mei J, Cao R, Liu X, Fu B, Ma M, et al. Integrative analysis identifies AKAP8L as an immunological and prognostic biomarker of pan-cancer. Aging 2023;15:8851–72. 10.18632/aging.205003.

[60] Yang L, Zhang H, Chen D, Ding P, Yuan Y, Zhang Y. EGFR and Ras regulate DDX59 during lung cancer development. Gene 2018;642:95–102. 10.1016/j.gene.2017.11.029.

[61] Renatino-Canevarolo R, Meads MB, Silva M, Sudalagunta PR, Cubitt C, De Avila G, et al. Dynamic Epigenetic Landscapes Define Multiple Myeloma Progression and Drug Resistance. Blood 2020;136:32–3. 10.1182/blood-2020-142872.

[62] Garofano F, Corsale AM, Biondo M, Romano A, Schmidt-Wolf I, Gullà AM, et al. Identification of CD320, SLC44A1 and TNFRSF13B as potential novel therapeutic targets for CAR T-cell therapy in multiple myeloma. Front Med (Lausanne) 2025;12:1737919. 10.3389/fmed.2025.1737919.

[63] Wise JF, Berkova Z, Mathur R, Zhu H, Braun FK, Tao R-H, et al. Nucleolin inhibits Fas ligand binding and suppresses Fas-mediated apoptosis in vivo via a surface nucleolin-Fas complex. Blood 2013;121:4729–39. 10.1182/blood-2012-12-471094.

[64] Hu J, Liu W, Yang T, Chen Y. PB2104 INHIBITION OF NUCLEOLIN REVERSED BORTEZOMIB RESISTANCE VIA THE REGULATION OF NF-κB PATHWAY IN MULTIPLE MYELOMA CELLS. Hemasphere 2019;3:948. 10.1097/01.HS9.0000566900.63412.df.

[65] Zhang J, Wang Z, Wang K, Xin D, Wang L, Fan Y, et al. Increased Expression of SRSF1 Predicts Poor Prognosis in Multiple Myeloma. J Oncol 2023;2023:9998927. 10.1155/2023/9998927.

[66] Liu N, Xie Z, Li H, Wang L. The numerous facets of 1q21+ in multiple myeloma: Pathogenesis, clinicopathological features, prognosis and clinical progress (Review). Oncol Lett 2024;27:258. 10.3892/ol.2024.14391.

[67] Shen X, Luo D, Yang X, Li Y, Lian F, Liu H, et al. Intercellular Adhesion Molecule 3: Structure, Cellular Functions, and Emerging Role in Human Diseases. J Cancer 2025;16:1563–74. 10.7150/jca.100612.

[68] Wang Y, Gao S, Chen L, Liu S, Ma J, Cao Z, et al. DUT enhances drug resistance to proteasome inhibitors via promoting mitochondrial function in multiple myeloma. Carcinogenesis 2022;43:1030–8. 10.1093/carcin/bgac071.

[69] Costacurta M, Vervoort SJ, Hogg SJ, Martin BP, Johnstone RW, Shortt J. Whole genome CRISPR screening identifies TOP2B as a potential target for IMiD sensitization in multiple myeloma. Haematologica 2021;106:2013–7. 10.3324/haematol.2020.265611.

[70] Heynen GJJE, Baumgartner F, Heider M, Patra U, Holz M, Braune J, et al. SUMOylation inhibition overcomes proteasome inhibitor resistance in multiple myeloma. Blood Adv 2023;7:469–81. 10.1182/bloodadvances.2022007875.

[71] Bam R, Khan S, Ling W, Randal SS, Li X, Barlogie B, et al. Primary myeloma interaction and growth in coculture with healthy donor hematopoietic bone marrow. BMC Cancer 2015;15:864. 10.1186/s12885-015-1892-7.

[72] Li L, Peng Y, Yuan Q, Sun J, Zhuang A, Bi X. Cathelicidin LL37 Promotes Osteogenic Differentiation in vitro and Bone Regeneration in vivo. Front Bioeng Biotechnol 2021;9:638494. 10.3389/fbioe.2021.638494.

[73] Liang J, Chen J, Ye Z, Bao D. Cathelicidin LL-37 improves bone metabolic balance in rats with ovariectomy-induced osteoporosis via the Wnt/beta-catenin pathway. Physiol Res 2022;71:369–77. 10.33549/physiolres.934820.

[74] Chen J, Lu S. Bioinformatics analysis of key genes, immune infiltration, and risk assessment in low bone mineral density among perimenopausal women: An observational study. Medicine 2024;103:e38695. 10.1097/MD.0000000000038695.

[75] Zhao K, Sun Y, Zhong S, Luo J-L. The multifaceted roles of cathepsins in immune and inflammatory responses: implications for cancer therapy, autoimmune diseases, and infectious diseases. Biomark Res 2024;12:165. 10.1186/s40364-024-00711-9.

[76] Gao G, Liu R, Wu D, Gao D, Lv Y, Xu X, et al. Risk score constructed with neutrophil extracellular traps-related genes predicts prognosis and immune microenvironment in multiple myeloma. Front Oncol 2024;14:1365460. 10.3389/fonc.2024.1365460.

[77] Cornish J, Callon KE, Naot D, Palmano KP, Banovic T, Bava U, et al. Lactoferrin is a potent regulator of bone cell activity and increases bone formation in vivo. Endocrinology 2004;145:4366–74. 10.1210/en.2003-1307.

[78] Naot D, Grey A, Reid IR, Cornish J. Lactoferrin--a novel bone growth factor. Clin Med Res 2005;3:93–101. 10.3121/cmr.3.2.93.

[79] Baranov M V., Revelo NH, Dingjan I, Maraspini R, ter Beest M, Honigmann A, et al. SWAP70 Organizes the Actin Cytoskeleton and Is Essential for Phagocytosis. Cell Rep 2016;17:1518–31. 10.1016/j.celrep.2016.10.021.

[80] Lennartsson A, Garwicz D, Lindmark A, Gullberg U. The proximal promoter of the human cathepsin G gene conferring myeloid-specific expression includes C/EBP, c-myb and PU.1 binding sites. Gene 2005;356:193–202. 10.1016/j.gene.2005.05.004.

[81] Elbahoty MH, Papineni B, Samant RS. Multiple myeloma: clinical characteristics, current therapies and emerging innovative treatments targeting ribosome biogenesis dynamics. Clin Exp Metastasis 2024;41:829–42. 10.1007/s10585-024-10305-2.

[82] Nikesitch N, Lee JM, Ling S, Roberts TL. Endoplasmic reticulum stress in the development of multiple myeloma and drug resistance. Clin Transl Immunology 2018;7. 10.1002/cti2.1007.

[83] Díaz-Tejedor A, Lorenzo-Mohamed M, Puig N, García-Sanz R, Mateos M-V, Garayoa M, et al. Immune System Alterations in Multiple Myeloma: Molecular Mechanisms and Therapeutic Strategies to Reverse Immunosuppression. Cancers (Basel) 2021;13. 10.3390/cancers13061353.

[84] Lee SJ, Borrello I. Role of the Immune Response in Disease Progression and Therapy in Multiple Myeloma. Cancer Treat Res 2016;169:207–25. 10.1007/978-3-319-40320-5_12.

[85] Eipper S, Steiner R, Lesner A, Sienczyk M, Palesch D, Halatsch M-E, et al. Lactoferrin Is an Allosteric Enhancer of the Proteolytic Activity of Cathepsin G. PLoS One 2016;11:e0151509. 10.1371/journal.pone.0151509.

[86] Chen Z, Xu X. Roles of nucleolin. Focus on cancer and anti-cancer therapy. Saudi Med J 2016;37:1312–8. 10.15537/smj.2016.12.15972.

[87] Tellier J, Nutt SL. Plasma cells: The programming of an antibody-secreting machine. Eur J Immunol 2019;49:30–7. 10.1002/eji.201847517.

[88] Zheng X, Wang Y, Duan H, Hou J, He S. A new NCL-targeting aptamer-drug conjugate as a promising therapy against esophageal cancer. J Nanobiotechnology 2025;23:52. 10.1186/s12951-025-03127-1.

[89] Granneman S, Gallagher JEG, Vogelzangs J, Horstman W, van Venrooij WJ, Baserga SJ, et al. The human Imp3 and Imp4 proteins form a ternary complex with hMpp10, which only interacts with the U3 snoRNA in 60-80S ribonucleoprotein complexes. Nucleic Acids Res 2003;31:1877–87. 10.1093/nar/gkg300.

[90] Eura Y, Ishihara N, Oka T, Mihara K. Identification of a novel protein that regulates mitochondrial fusion by modulating mitofusin (Mfn) protein function. J Cell Sci 2006;119:4913–25. 10.1242/jcs.03253.

[91] Watanabe Y, Tamura Y, Kakuta C, Watanabe S, Endo T. Structural basis for interorganelle phospholipid transport mediated by VAT-1. J Biol Chem 2020;295:3257–68. 10.1074/jbc.RA119.011019.

[92] Junker M, Rapoport TA. Involvement of VAT-1 in Phosphatidylserine Transfer from the Endoplasmic Reticulum to Mitochondria. Traffic 2015;16:1306–17. 10.1111/tra.12336.

[93] Perez-Riverol Y, Bandla C, Kundu DJ, Kamatchinathan S, Bai J, Hewapathirana S, et al. The PRIDE database at 20 years: 2025 update. Nucleic Acids Res 2025;53:D543–53. 10.1093/nar/gkae1011.

