## Supplementary figures and images for "Integrated Proteomic and Network Analysis Reveals Dysregulated Pathways and Candidate Proteins in Multiple Myeloma Progression"

### Additional File 5

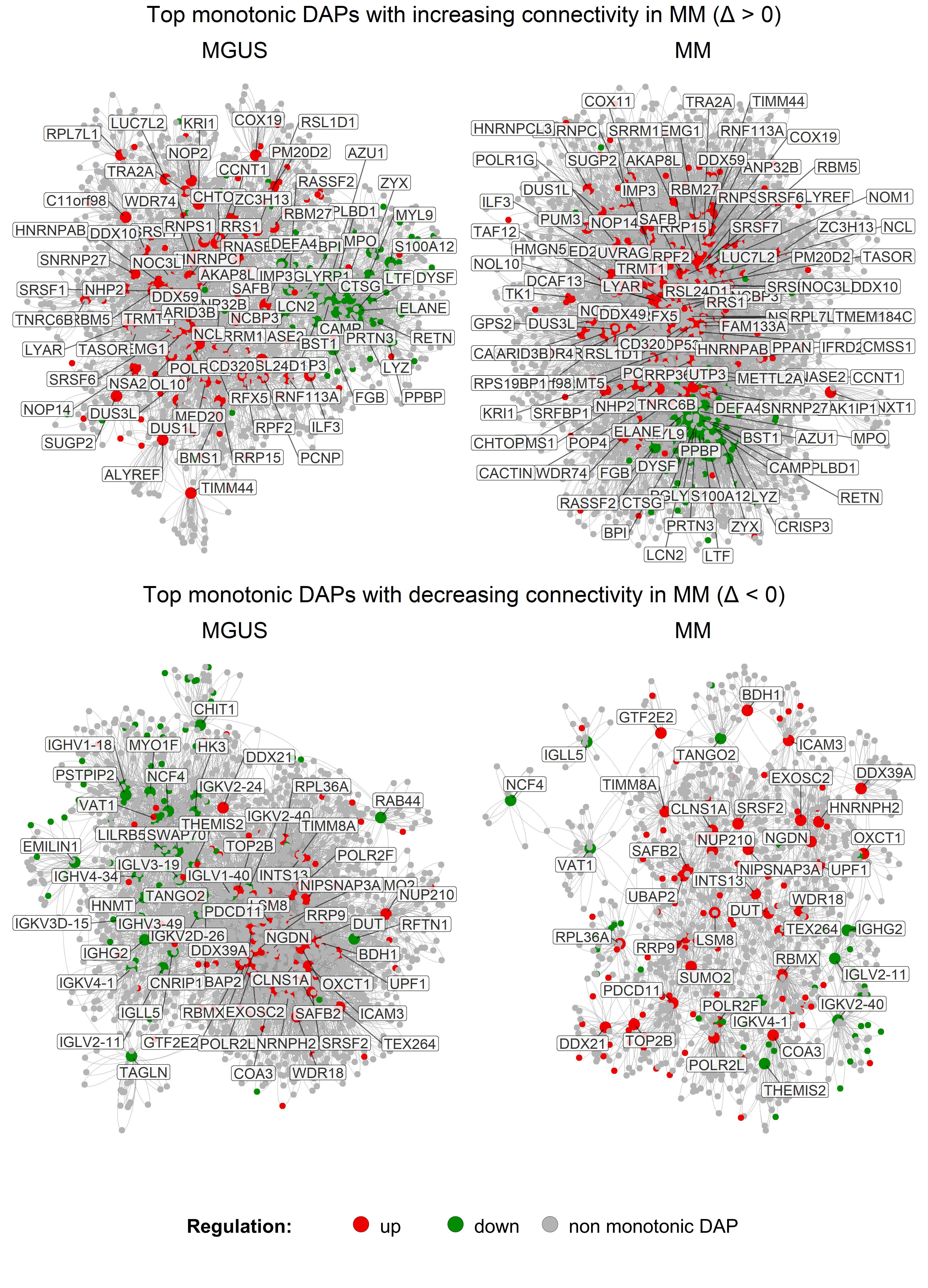
