## Additional File 7 for "Integrated Proteomic and Network Analysis Reveals Dysregulated Pathways and Candidate Proteins in Multiple Myeloma Progression"

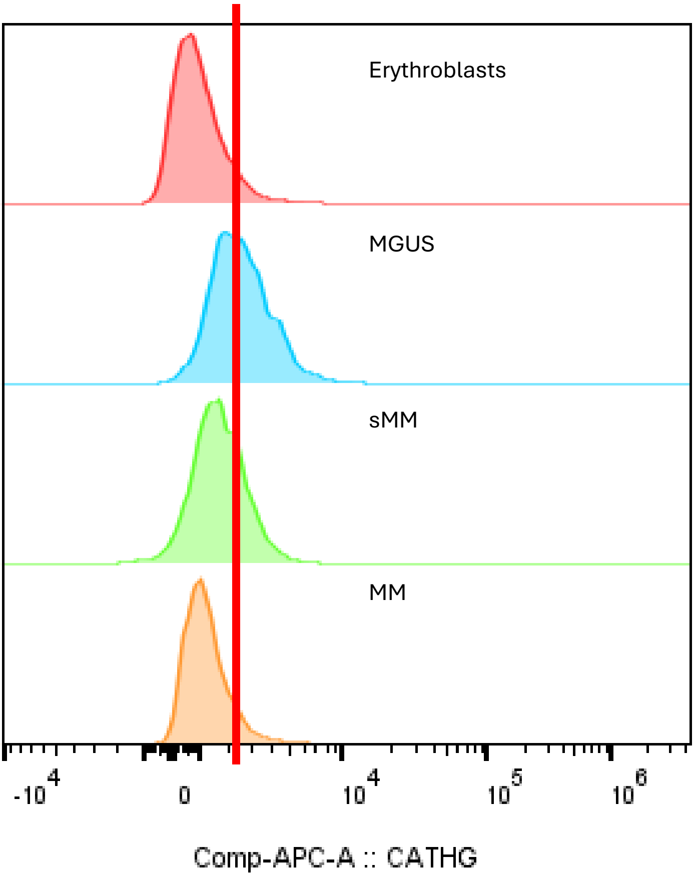


| | Subset Name | Median : Comp-APC-A |
|---|---|---|
| | Erythroblasts | 793 |
| | Clonal plasma cells | 2441 |
| | Clonal plasma cells | 1655 |
| | Clonal plasma cells | 1098 |
